# Complex structural variation, phylogeny, and disease associations of the mucin pangenome

**DOI:** 10.64898/2026.07.01.26356476

**Authors:** Elizabeth G. Plender, Timofey Prodanov, Jiadong Lin, Isaac Wong, Julie Wertz, William W. Gordon, Michael J. Bamshad, Katherine M. Munson, Wanda K. O’Neal, Jesse D. Bloom, Human Pangenome Reference Consortium, Tobias Marschall, Evan E. Eichler

## Abstract

Mucins are large glycoproteins that provide hydration and barrier function to epithelial tissues. Although genetically heterogeneous, all mucins harbor a large exon composed of variable number tandem repeats (VNTRs). Short-read sequencing has limited our understanding of mucin VNTR diversity and makes disease association studies challenging. We leverage 296 long-read phased genome assemblies to characterize 14 mucin family members, achieving ≥97% accuracy across 572 haplotypes. Phylogenetic haplogroup analysis reveals extraordinary structural heterozygosity, with *MUC4* harboring the greatest allelic diversity (n=240 distinct lengths) and *MUC12* the greatest size range (Δ = 55,233 bp; 23,080 amino acids). Ten mucins show significant population stratification (pFDR < 0.05). At the *MUC4/MUC20* locus, we characterize higher-order structural variation, including a recurrent inversion, copy number variation, and interlocus gene conversion. Optimized genotyping achieves ≥95% haplogroup concordance across 10 loci. We apply this to 4,637 deeply phenotyped cystic fibrosis patients and identify a significant association between short *MUC1* VNTRs and severe disease (p=0.0056), demonstrating the pangenome’s utility for complex locus genotyping and disease discovery.

## INTRODUCTION

Mucus is an essential secretion that maintains epithelial homeostasis. It hydrates the airways, gastrointestinal tract, and reproductive system and provides a defensive role against inhaled or ingested environmental pathogens and particulates^1^. The biochemical and physical barrier properties of mucus are largely attributed to mucins, a highly diverse family of glycoproteins^2^. These massive proteins harbor one or more repeat domains rich in serine and threonine that serve as scaffolds for dense arrays of O-linked glycans, thereby mediating water retention and interactions with sugars on the surface of pathogens^3^. They partition into two functional classes: secreted and tethered mucins. The secreted, gel-forming mucins polymerize via conserved cystine-rich D-domains and form the visible mucus layer^4^, while the tethered mucins anchor to the apical surface of epithelial cells and form a glycocalyx that provides a secondary layer of hydration and defense^5^.

All canonical mucins share a common architectural feature—their repeat protein domains are encoded by a large exon composed of variable number tandem repeats (VNTRs). Variation in the length of these repeats and their sequence composition may modulate the number and spacing of glycosylation sites, thereby impacting biophysical properties of the mucus like gel viscosity, barrier thickness, mucociliary clearance dynamics, and pathogen binding capacity^3^. Additionally, VNTRs mutate at several orders of magnitude higher than unique sequence through mechanisms like replication slippage and unequal crossing over, producing extensive length and sequence variation^6^. The mucins are an ancient gene family that have likely evolved under particular selective pressures, including sustained host–pathogen conflict^7^. Combined, these sequence features contribute to extensive length polymorphism in these genes across human populations.

Variation at functionally critical loci often shapes disease risk, and mucins are no exception. Genome-wide association studies (GWAS) have implicated the mucins in a broad spectrum of epithelial diseases with mucus pathology, including chronic airway conditions like asthma^8^ and cystic fibrosis^9^, gastrointestinal disorders like inflammatory bowel disease^10^, and many cancers^11^. The contribution of mucin genetic variation to these phenotypes has been challenging to resolve due to the complexity of these loci. Where mucin variation has been resolvable, it has yielded strong disease signals, exemplified by the *MUC5B* promoter polymorphism in idiopathic pulmonary fibrosis^12^. For nearly all other cases, however, limitations in direct genotyping of the VNTR have made association testing challenging. Causal variation, instead, is more likely tagged by surrounding single-nucleotide polymorphisms (SNPs) rather than directly genotyped, with the underlying mucin haplotypes remaining uncharacterized. Mucin VNTRs are widely suspected to account for a meaningful share of the missing heritability of epithelial disease^13^, but resolving this contribution has required tools and sequence resolution of the VNTRs that did not exist until recently.

The fundamental obstacle to studying the genetic contribution of mucins to disease has been limitations of short-read sequencing. Short reads (∼250 bp) cannot unambiguously resolve mucin VNTRs because they are shorter than most mucin repeat arrays, leading to multi-mapping and missassembly^14^. Until recently, mucin loci in human reference assemblies contained errors and gaps. Consequently, imputation panels built from short-read data systematically excluded variation at these genes^15^; however, long-read sequencing for haplotype-resolved pangenome assemblies now overcomes these limitations. Reads from current long-read platforms routinely span entire mucin VNTR arrays, enabling complete reconstruction of individual haplotypes^16^. Reference-building consortia like the Human Pangenome Reference Consortium (HPRC)^17^ and the Human Genome Structural Variation Consortium (HGSVC)^15^ further provide phased, assembly-based references from genetically diverse individuals, eliminating bias in mapping to a single linear genome^18^. Together, these advances make it possible to accurately characterize mucin genetic variation at scale.

We previously characterized structural genetic diversity for the secreted mucins *MUC5AC* and *MUC5B* using ∼100 long-read genome assemblies from the HPRC and HGSVC, demonstrating the feasibility of long-read assembly at these loci^19^. Here, we expand upon that work to establish a general strategy for assessing 14 members of the canonical mucin gene family. We assemble and evaluate these secreted and tethered mucins across a larger and more genetically diverse sample set (296 genomes), enabling cross-family comparisons of mucin sequence architecture and evolution. This expanded haplotype catalog improves the accuracy of mucin genotyping from short-read data and facilitates downstream association testing. As a proof of principle, we apply this framework to a cystic fibrosis cohort and show that mucin haplotypes are candidate modifiers for clinically relevant phenotypes. The strategies outlined here extend to a broad range of epithelial diseases, as well as other biomedically relevant multiallelic loci in the human genome.

## RESULTS

### Mucin gene models and pangenome assembly

The mucin gene family encodes functionally related glycoproteins involved in mucosal protection and signaling^20^, though these genes do not represent a single ancestrally related family^21^. To systematically assess variation across mucin genes, we used the CHM13 genome assembly^22^ to annotate gene models and open reading frames (ORFs), overcoming the incomplete assembly, collapse, or gaps that characterize mucin VNTRs in standard reference genomes. Loci were selected based on canonical mucin nomenclature and annotations for VNTR status/mucin subclass (secreted vs. tethered) from Chaturvedi et al.^23^ Two additional mucins, *MUC21* and *MUC22*, have since been added to the gene family^24,25^. We excluded *MUC3B* (no CHM13^22,26^ gene annotation) and *MUC13* (no VNTR-containing exon in CHM13), thereby retaining 15 loci for further analyses: *MUC1/2/3A/4/5AC/5B/6/7/12/16/17/19/20/21/22.* These genes are distributed across eight chromosomes, with notable clustering on chromosome 3 (*MUC4* and *MUC20*), chromosome 6 (*MUC21* and *MUC22*), chromosome 7 (*MUC3A*, *MUC12*, and *MUC17*), and chromosome 11 (*MUC6*, *MUC2*, *MUC5AC*, and *MUC5B*).

The other four mucins are dispersed as single genes on chromosome 1 (*MUC1*), chromosome 4 (*MUC7*), chromosome 12 (*MUC19*), and chromosome 19 (*MUC16*). For *MUC1*, *MUC3A*, *MUC4*, *MUC12*, and *MUC19*, GRCh38-to-CHM13 liftover^27^ produced segmented VNTR exons, necessitating manual curation to maintain ORFs across the repeat sequences (Figure 1a, Supplementary Table 1). All loci except *MUC19* maintained an ORF in CHM13. Although Zhu et al.^28^ showed that the *MUC19* central exon lacks intron interruption in the longest isoform, we identified an indel in CHM13 that prematurely disrupts the ORF. We found no complete ORF for *MUC19* in any of our long-read haplotypes and therefore excluded this locus from further analysis.

**Figure 1.**
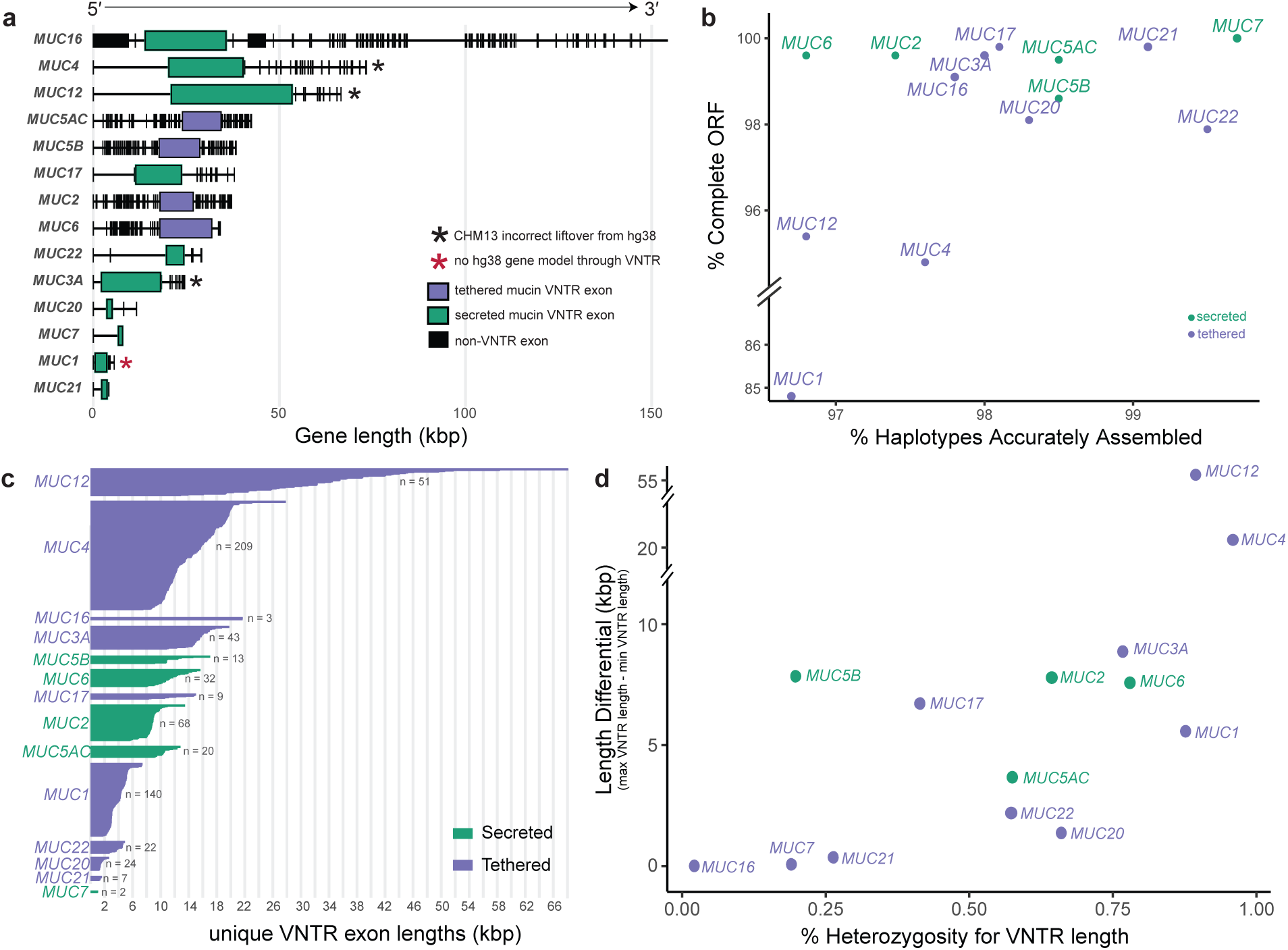
The tethered mucins are more variable than the secreted mucins in humans. **a,** CHM13 gene model annotations for the secreted and tethered mucins. Black asterisks (*) represent loci with incorrect liftover between hg38 and CHM13, requiring manual curation to complete the VNTR exon. Red asterisks indicate loci with incomplete annotation in hg38 and CHM13, requiring manual curation to complete the VNTR exon. **b,** Scatterplot of locus assembly quality (via Flagger) and open reading frame (ORF) maintenance across haplotypes in the canonical secreted and tethered mucins chosen for inclusion in this study. Loci were chosen based on presence of a coding VNTR and complete ORF across most haplotypes (*MUC19* excluded due to the lack of an ORF for all long-read haplotypes). **c,** Unique VNTR exon length alleles per secreted and tethered mucin locus. **d,** Scatterplot of percentage of length heterozygosity (irrespective of VNTR sequence variation) across haplotypes versus the length differential between alleles (largest allele minus smallest allele) across mucin loci.

For the remaining 14 loci, we analyzed long-read genome assemblies from 296 diverse humans from the HPRC (release 1: n = 1, release 2: n = 231) and HGSVC (phase 2: n = 4, phase 3: n = 60) sample sets (Supplementary Table 2). Compared to our previous analysis of *MUC5AC* and *MUC5B*^19^, this sample set represents a ∼3-fold expansion. Based on Flagger and HMM-Flagger^29^, we estimate that 96.8%–99.7% of haplotypes are accurately assembled depending on the locus (Figure 1b). Haplotype mapping of the longest isoforms yielded 94.8%–100% ORF maintenance across loci. In general, ORF completeness correlates inversely with structural heterozygosity (Figure 1c-d) except for *MUC1* (84.8%). This is likely attributable to higher rate of sequencing errors in the GC-rich homopolymer VNTRs of *MUC1*^30,31^.

### Structural heterozygosity of mucin VNTRs

Genome-wide analyses indicate that mucins are among the most structurally heterozygous repeats in humans, with 4 out of the top 10 most variable loci containing coding short tandem repeats (STRs) or VNTRs being mucins (*MUC4*, *MUC12*, *MUC6*, and *MUC2*)^17^. When examining unique alleles per locus for VNTR exon size, the secreted mucins are more similar to one another compared to the tethered mucins (Figure 1b-c). Among the secreted mucins, *MUC2* exhibits the highest allelic diversity (n = 68), yet this remains modest relative to the tethered mucins *MUC1* (n = 140) and *MUC4* (n = 209). While *MUC4* is both extensively heterozygous and features long alleles (longest allele at 27,843 bp), *MUC1* features much smaller alleles (longest allele at 7,408 bp). *MUC16* harbors the third largest VNTR size yet only has three unique alleles that differ by 3-6 base pairs. *MUC12* harbors a modest number of unique alleles (n = 51); however, this locus has a sizable length differential between the longest and shortest alleles (Figure 1d). The shortest predicted protein for *MUC12* among our haplotypes is 4,677 amino acids (aa) in length, while the longest is 23,080 aa. This indicates that MUC12 is one of the largest proteins in the human proteome^32^ and contains the largest exon in the human genome^33^ (far longer than the 27,303 bp exon in *GRIN2B*).

To assess how VNTR variation is distributed across human populations, we first examined existing 1000 Genomes Project (1KGP) superpopulation annotations^34^ for the 296 HPRC and HGSVC haplotypes—100 African (AFR), 52 Admixed American (AMR), 61 East Asian (EAS), 39 European (EUR), and 44 South Asian (SAS). We then compared these annotations with continuous local ancestry coordinates computed at each mucin locus using PCLAI (Point Cloud Local Ancestry Inference^35^) for HPRC release 2 haplotypes (n = 231). AMR haplotypes exhibited the greatest local-ancestry admixture and did not form a discrete cluster in PCLAI space (consistent with their admixed designation) while the remaining four superpopulations were clearly resolved (Extended Data Figure 1). Across the 14 loci, superpopulation explained 85–89% of variance in PCLAI coordinates (PERMANOVA R²), rising to 94–97% when AMR haplotypes were excluded. We therefore extracted the four PCLAI-derived ancestry clusters for HPRC haplotypes at each locus, which are denoted as AFR ancestry, EUR ancestry, SAS ancestry, and EAS/AMR ancestry, to more faithfully reflect local ancestry compared to broad superpopulation labels from geographic criteria. These groupings provide sufficient per-group sample sizes for well-powered comparisons. Given the biomedical relevance of mucin coding structural variation, preserving this ancestry stratification was essential to resolving population-differentiated patterns that may carry functional consequences in the mucins.

Tethered mucins show overall greater population stratification, with *MUC4* carrying the highest proportion of population-specific alleles (∼67%) private to AFR ancestry (Figure 2a). In addition, *MUC4* exhibits the greatest ancestry cluster variance of any locus (Figure 2b). Among the secreted gel-forming mucins, private alleles exist across all loci; however, they manifest as directional size differences rather than extreme variance. For example, AFR ancestry alleles carry the smallest VNTRs at *MUC6* and *MUC5AC*, EAS/AMR ancestry alleles carry the largest *MUC5AC* VNTRs, and AFR ancestry alleles harbor the largest *MUC5B* alleles—differences that remain statistically significant after multiple testing correction (Extended Data Figure 1). Even shared genomic regions on chromosome 11 (Figure 2c, Supplementary Figure 1) have diverged radically in recent human evolution. *MUC17* is unique in that it harbors few private alleles yet ranks as the third most population-stratified mucin, indicating that its alleles are broadly shared across populations but at markedly different allele frequencies.

**Figure 2.**
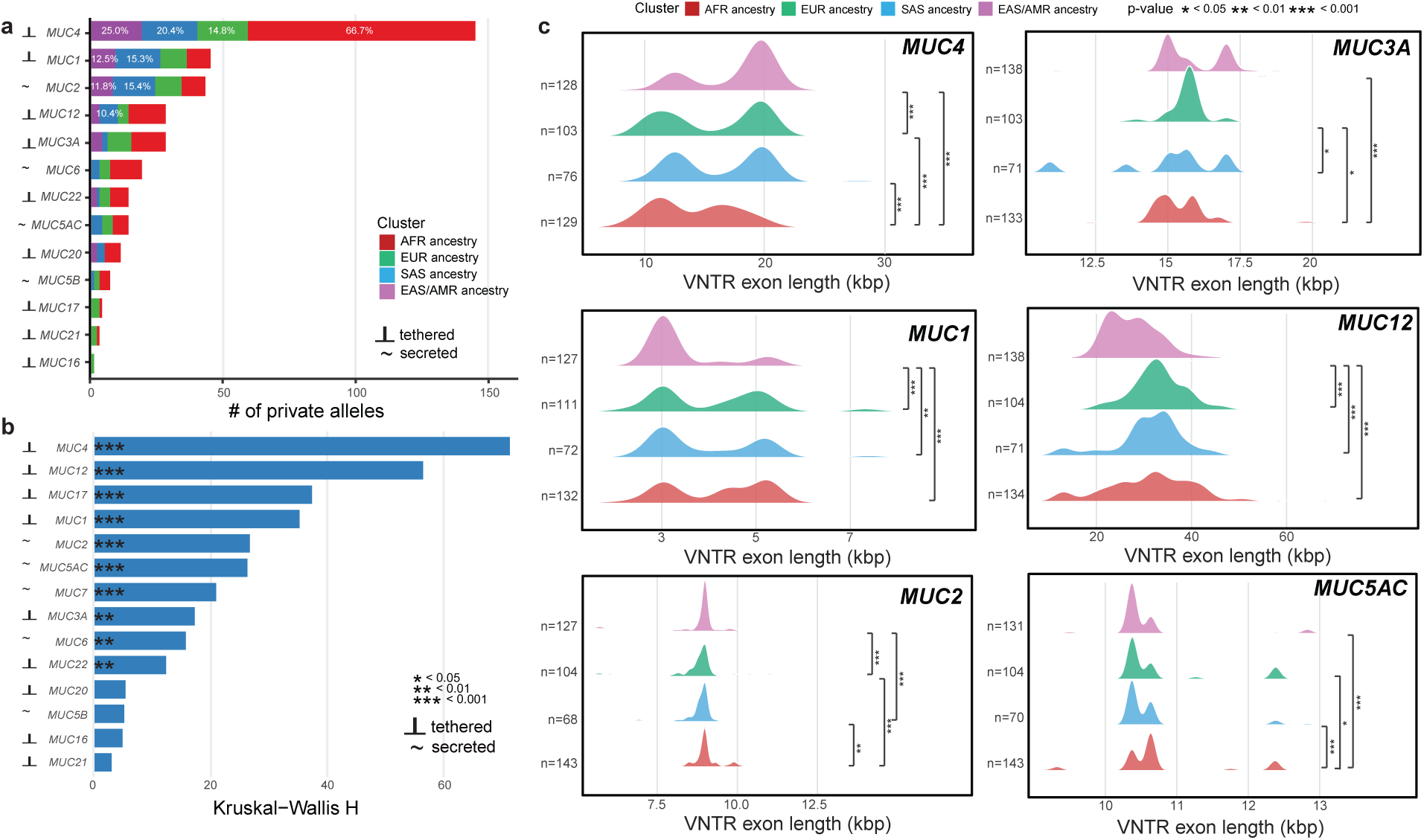
Population stratification of the secreted and tethered mucins in humans. **a,** Number of alleles private to single ancestry clusters across mucin loci (excluding *MUC7* due to presence of all unique alleles across clusters). Percentages are indicative of the number of alleles per ancestry cluster relative to a given mucin’s distribution and only those ≥10% are shown. **b,** Kruskal-Wallis test of ancestry cluster variance (all four clusters) in mucin VNTR exon size (bp) frequencies (H statistic, df = 4, FDR adjusted). A higher H statistic indicates that a mucin has strong variance in VNTR length across ancestry clusters. **c,** Ridge plots of VNTR length distributions per ancestry cluster at the top loci for private alleles (*MUC4*, *MUC1*, *MUC2*, *MUC12*, and *MUC3A*; Figure 1a) and population variance (*MUC4*, *MUC1*, *MUC2*, *MUC12*, *MUC3A*, and *MUC5AC*; Figure 1b). P-values correspond to pairwise Wilcoxon rank-sum tests (FDR-adjusted).

### Mucin VNTR protein domain and motif variation

In addition to length polymorphisms, mucin VNTRs can display protein domain and amino acid compositional variation that can have functional consequences^36,37^. The VNTR exons of the secreted gel-forming mucins *MUC2*, *MUC5AC*, and *MUC5B*, for example, encode both tandem repeat and cysteine-rich (cys) domains (Extended Data Figure 2). By expanding our reference haplotype set, we uncovered seven additional *MUC5AC* protein variants, including a long variant private to EAS/AMR ancestry haplotypes (6,473 aa) that feature an additional tandem repeat domain (6 total). At *MUC5B*, five additional variants were identified, the largest of which (7,821 aa) contains eight tandem repeat domains. This also exceeds the previous maximum by one domain. In contrast, *MUC2* contains two invariant cys domains and two tandem repeat domains that are fixed in copy number across haplotypes, with only the second repeat domain varying in length.

Serine and threonine residues are the primary sites of O-glycosylation in mucins; therefore, we examined their composition across haplotypes (Figure 3a, see Supplementary Table 3 for full list of proteins). Each mucin occupies a distinct and well-separated rank in S/T composition, ranging from ∼25% in *MUC1* to ∼52% in *MUC3A*, with little variation between haplotypes within a locus. This is consistent with the notion that S/T content is tightly constrained. Notably, this ranking does not follow the secreted/tethered distinction; instead, *MUC1*, and *MUC20*, which show the broadest expression across mucins in GTEx epithelial tissues (p = 0.014, Wilcoxon rank-sum), cluster at the lower end of the S/T distribution relative to more tissue-restricted mucins (both p<0.0001, Wilcoxon rank-sum). Additionally, we found that proline content was significantly negatively correlated with S/T content across all 14 mucin VNTR domains (Spearman’s ρ = −0.63, p = 0.017), suggesting a compositional tradeoff between proline-induced backbone rigidity and O-glycosylation propensity (Extended Data Figure 3).

**Figure 3.**
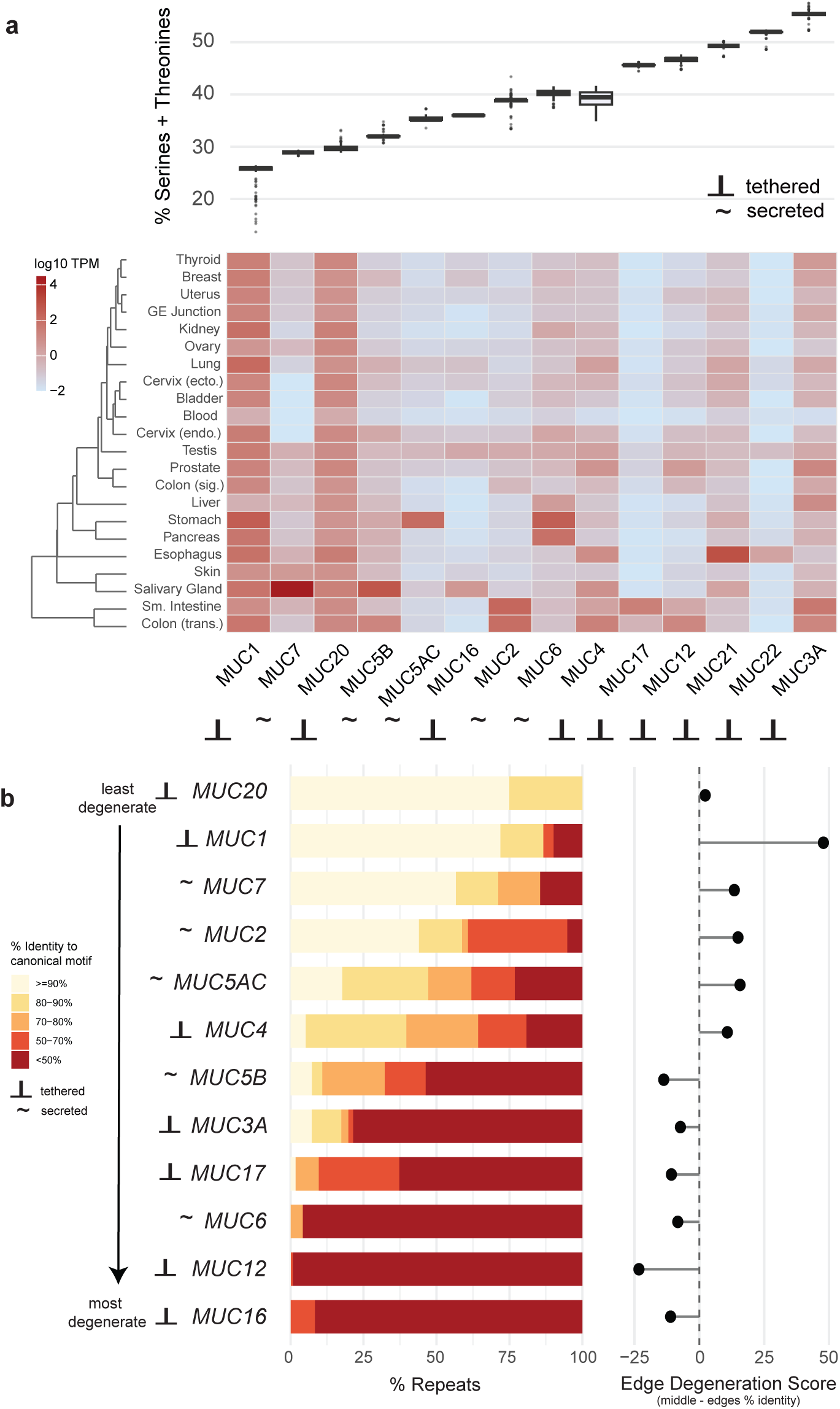
VNTR motif composition in the secreted and tethered mucins. **a,** Percent composition of serines and threonines in haplotypes with complete ORFs across mucins. Heatmap depicts log10 TPM (transcripts per million) from epithelial tissue RNA sequencing data in GTEx. **b,** Percentage of repeats containing sequence divergence from the canonical motif per mucin across haplotypes and edge degeneration scores. Score calculated as average percent identity across all edges minus average percent identity across all middle repeats (edge # = 10% of median repeat count). Positive values indicate greater degeneration at array boundaries.

We next characterized VNTR amino acid motif variation across loci to assess repeat degeneration and to establish canonical motifs for proteomics applications (Figure 3b, Extended Data Figure 4). Comparing our haplotype-derived canonical (most frequent) motifs to those reported by Chaturvedi et al.^23^, we found discordance at eight loci (*MUC2*, *MUC4*, *MUC5B*, *MUC6*, *MUC7*, *MUC12*, *MUC16*, and *MUC17*). Approximately half of the mucins (*MUC1*, *MUC2*, *MUC4*, *MUC5AC*, *MUC7*, and *MUC20*) feature more motif degeneration at array boundaries than in internal repeats, consistent with reduced homogenization of terminal repeats in tandem repeat evolution^38^. While *MUC16* harbors very little length variation, it does harbor the most degenerate VNTR, suggesting that single-nucleotide variation is under comparatively relaxed selective constraint at this locus.

### Evolution of the mucin VNTRs

Previous studies have indicated that the four secreted mucins mapping to chr11p15 (*MUC6*, *MUC2*, *MUC5AC*, and *MUC5B*) are situated in a recombination-rich region of the genome^39^. We tested this region and other mucin loci relative to genome-wide rates using recombination maps constructed with CHM13 for five populations representative of continental groups in the 1KGP samples^40^. These included Yoruba in Nigeria (YRI), Utah residents from North and West Europe (CEU), Han Chinese in Beijing (CHB), Gujarati Indians in Houston (GIH), and Peruvians in Lima (PEL). Loci closer to telomeres showed higher recombination rates (chr3q29, chr11p15, chr19p13, and chr6p21), though none were outliers relative to the genome-wide distribution (Extended Data Figure 5). Conversely, chr1q21 (*MUC1*) in YRI and chr4q13 (*MUC7*) in CHB showed diminished recombination.

We compared human mucin gene structures to eight recently sequenced and assembled nonhuman primate (NHP) genomes representing ∼35 million years of primate evolution^41–43^. While the sample size for each of these species is too few to comment on intraspecies variation, this general comparative analysis allows us to comment on overall trends in the size of mucin VNTRs since human divergence. *MUC1*, *MUC4*, and *MUC22* exhibit longer VNTRs in humans relative to NHPs, whereas *MUC5AC*, *MUC5B*, and *MUC6* trend shorter, indicating that some mucins display lineage-specific expansions and contractions rather than a consistent directional shift (Figure 4a).

**Figure 4.**
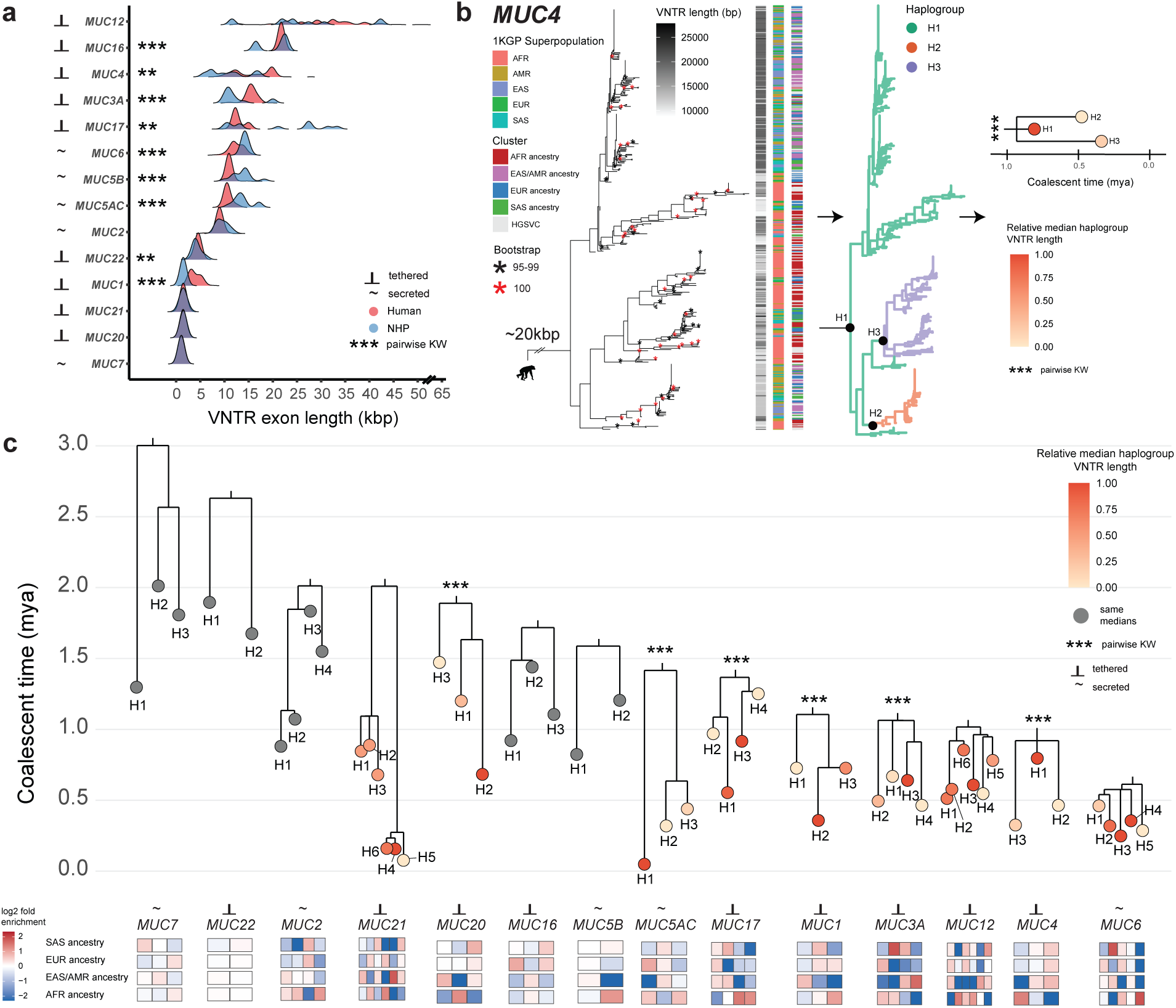
Comparative evolution and phylogenetic history of mucin loci. **a,** Ridge plot comparing human and nonhuman primate (NHP) VNTR exon allele sizes for mucin loci. One individual each for chimpanzee, bonobo, gorilla, Sumatran orangutan, Bornean orangutan, macaque, and marmoset included. KW indicates pairwise Kruskal-Wallis test. **b,** Schematic of mucin phylogenetic history reconstruction, haplogroup prediction, and tree simplification, using *MUC4* as an example. Phylogenetic trees for each locus were constructed with neutral sequences (non-repetitive intronic flank sequence) from all Flagger-verified human haplotypes and two chimpanzee haplotypes for outgrouping (see Supplementary Figures 5-8 for full trees). Haplogroups were determined via a threshold scanning approach that integrates tree topology, branch lengths, VNTR lengths, and bootstrap information for clustering. The simplified haplogroup tree was constructed by cutting the tree at the MRCA per group and dating was performed using chimpanzee outgrouping (6.4 million years ago [mya] divergence estimate). **c,** Simplified depictions of human mucin phylogenetic histories, subsetted to representative nodes for allele haplogroups. Tip colors represent median VNTR length per haplogroup. Significant pairwise Wilcoxon rank-sum tests with Benjamini-Hochberg correction denoted with ***. Heatmap below trees represents log2fold enrichment of ancestry cluster representations within mucin-specific haplogroups.

To investigate mucin evolution within the human lineage, we constructed phylogenetic trees for each locus using neutrally evolving intronic and flanking sequence (Methods, Figure 4b, Supplementary Figures 2-5) and mapped VNTR exon lengths and ancestry cluster designations onto the resulting topologies. Given the substantial structural and compositional heterogeneity across loci, we sought to reduce haplotypic complexity to improve power for future disease association studies. We therefore applied a systematic unsupervised clustering framework based on phylogenetic distances to group haplotypes into haplogroups, enabling both simplification of locus-specific trees and estimation of coalescent times using chimpanzee as an outgroup (6.4 million years ago [mya] divergence; Figure 4c). While *MUC7*, *MUC22*, *MUC2*, *MUC16*, and *MUC5B* showed no significant differences in median VNTR length among haplogroups, six loci (*MUC20*, *MUC5AC*, *MUC17*, *MUC1*, *MUC3A*, and *MUC4*) contained haplogroups with significantly different median VNTR lengths, indicating that short, intermediate, and long VNTR alleles have arisen and persisted over evolutionary timescales and can therefore be genetically tracked through haplogroup structure.

### Complex structural variation in chr3q29 and *MUC20* copy number variability

*MUC4* and *MUC20* map to chr3q29, a genomic region with complex patterns of structural variation. Both loci are flanked by segmental duplications that mediate recurrent inversions and copy number variability^44,45^. We distinguish 20 distinct structural configurations across human haplotypes based on *MUC4* and *MUC20* orientation and partial and complete gene duplications of the loci (Figure 5a). The inversion is strongly population-stratified, with AFR ancestry haplotypes carrying the inverted configuration at roughly 50% frequency compared to 25–30% in other superpopulations (Extended Data Figure 6). Non-inverted haplotypes associate with longer *MUC4* alleles (p adjusted < 0.001), while *MUC20* VNTR length does not correlate with inversion status (Extended Data Figure 6. Approximately 30% of haplotypes carry two or more complete *MUC20* paralogs and there is evidence that these additional copies are expressed^44^. Phylogenetic reconstruction of *MUC20* (Methods) ancestral and complete additional copies suggests that the duplication has occurred recurrently (Figure 5b) and has been subjected to interlocus gene conversion based on three complementary methods (GENECONV, Bootscan, and 3Seq^46,47^; p = 4.0×10⁻³¹ to 5.0×10⁻²; Supplementary Table 4).

**Figure 5.**
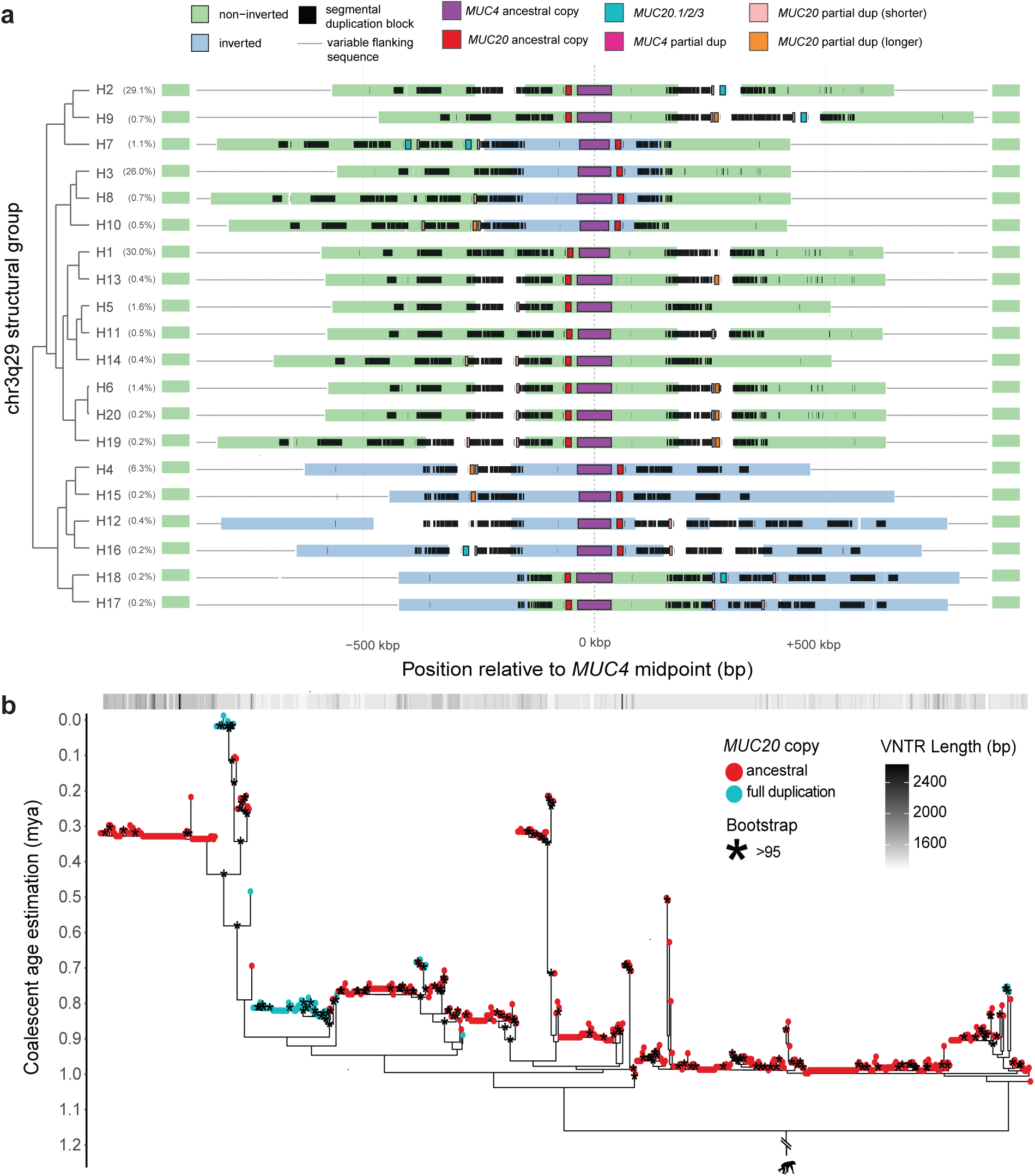
*MUC4* and *MUC20* are within a hypervariable region of chr3q29 flanked by segmental duplications. **a,** Unique structural configurations of chr3q29 across 570 haplotypes relative to a reference haplotype for the most common configuration (H1). Colored structures with arrows indicate inversion orientation (blue and green) and black boxes indicate DupMasker segmental duplication tracks. White tracks without orientation signify additional sequence relative to reference haplotype. Percentages represent configuration frequency across haplotypes and tree represents hierarchical clustering of configuration types. *MUC20* ancestral copy was inferred based on closest complete copy to the *MUC4* ancestral gene. *MUC20.1/2/3* denotes additional full copies distinct from the ancestral copy. **b,** Phylogenetic tree of *MUC20* ancestral and duplicate copies with mapped VNTR length distribution (constructed from all exon/intron sequences, excluding the central VNTR exon, ∼19 kbp total input sequence). To compare with the phylogenetic tree constructed without interlocus gene conversion tracks, see Supplementary Figure 6.

### GWAS risk allele mapping to pangenome haplotypes

Mucins are implicated in numerous disease associations through GWAS^9,30,48^, yet a fundamental limitation of these studies is that the identified tag SNPs have not been linked to variation in the VNTR^49^—the region known to encode, at least in part, mucin functionality. Reported variants may instead tag a haplotype on which a distinct causal variant is co-localized. Mucin structural variation likely represents an underappreciated source of disease risk, predominantly because VNTR alleles are poorly captured by SNP arrays^13^. To assess whether coding VNTRs could underlie these associations, we mapped GWAS risk alleles to pangenome haplotypes and tested for correlations between SNP genotype and VNTR length.

After filtering and processing (Methods, Supplementary Table 5), we identified five loci that had one or more SNPs significantly correlated with VNTR length (FDR adjusted): *MUC1*, *MUC4*, *MUC5AC*, *MUC6*, and *MUC22* (Figure 6a). *MUC4*, for example, had two SNPs supporting associations between longer alleles and COVID-19 severity, while multiple SNPs tagged shorter alleles with cystic fibrosis severity/bicuspid aortic valve (congenital heart defect). Similarly, *MUC1* had one SNP associating short alleles with lower lung function (FEV1). At *MUC5AC*, four SNPs were associated with longer alleles and gastric cancer risk. While these associations require further functional investigation, they raise the possibility that VNTR length variation at these loci may be a contributing factor to disease risk that has been overlooked in standard GWAS approaches.

**Figure 6.**
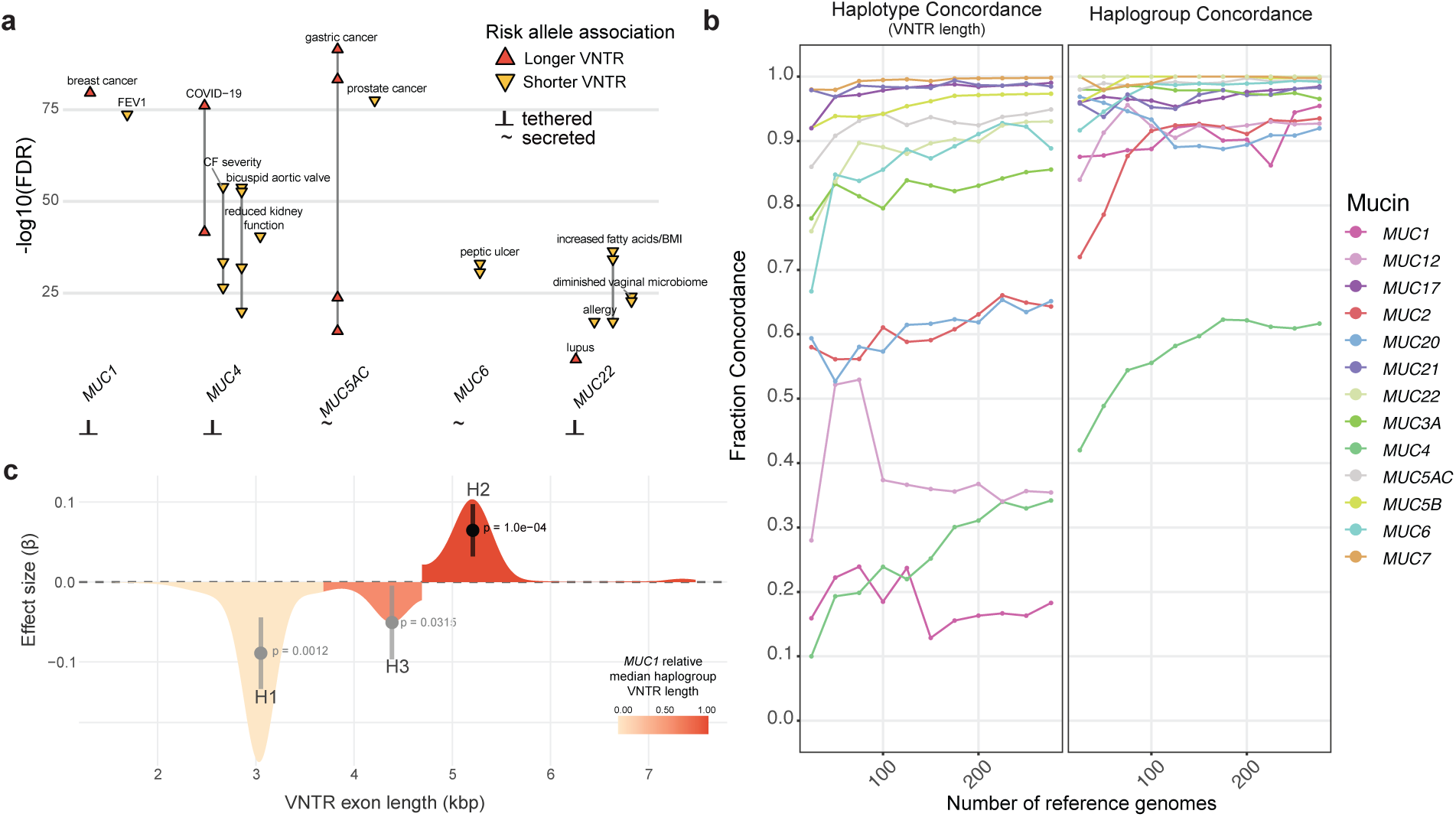
GWAS SNP mapping and Locityper genotyping support *MUC4* and *MUC1* as modulator loci of severe cystic fibrosis. **a,** SNPs with risk variants on the same VNTR length haplotypes from the HPRC/HGSVC genomes, grouped by nearest mucin gene. Lines connect SNPs with the same phenotype and VNTR length correlation. **b,** Locityper leave-one-out genotyping concordance with progressive increases in the number of reference genomes. **c,** Association testing (dosage model) of cystic fibrosis whole-genome sequencing data with Locityper-predicted *MUC1* genotype. P-values are Bonferroni adjusted for multiple hypothesis testing.

### Locityper mucin genotyping accuracy in short-read data

While mapping GWAS risk alleles to long-read pangenome haplotypes enables indirect inference of VNTR associations, direct genotyping of mucin VNTRs from short-read sequencing data would allow association testing in the large cohorts where GWAS are typically performed. To first assess the feasibility of direct mucin VNTR genotyping from short-read sequencing data, we applied Locityper v1.4.5^50^ to 276 genomes with paired long- and short-read data from the HPRC/HGSVC and 1KGP, using approximately three times more reference haplotypes than previous studies^19,50^. Leave-one-out cross-validation revealed substantial variation in genotyping accuracy across loci (Figure 6b, see Methods). At the haplotype level, only five mucins achieved ≥95% concordance (*MUC7*, *MUC17*, *MUC21*, *MUC5B*, and *MUC5AC*) and the most structurally complex loci showed poor performance, such as *MUC4* (∼35%), *MUC1* (∼17%), and *MUC12* (∼14%). In lieu of haplotype-specific genotyping, we considered our reduced complexity analysis of haplogroups. We found that accuracy improved at the haplogroup level, with *MUC1* reaching ∼93% concordance, though *MUC4* and *MUC12* remained challenging to genotype reliably. We also tested five region-specific weighting schemes to prioritize different genomic intervals during genotyping, but none improved accuracy beyond standard uniform weighting (Extended Data Figure 7). Subsampling analysis revealed that increasing reference panel size would benefit most loci, but several showed evidence of plateauing (Extended Data Figure 8, see Methods). For haplotype-level genotyping, *MUC1*, *MUC4*, and *MUC12* reached saturation at 36%, 37%, and 78% concordance, respectively, suggesting fundamental limitations due to extensive allelic diversity; however, haplogroup-level genotyping showed potential for improvement with larger reference panels, requiring an estimated 342, 1,105, and 278 additional genomes, respectively, to achieve 95% concordance (see Methods for information about model used).

### Mucin genotyping and association testing in a cystic fibrosis cohort

Based on our genetic characterization of the mucin pangenome, we sought to test whether new associations could be discovered for cystic fibrosis, where disease severity is frequently linked to mucus dysfunction^51^. To evaluate the potential clinical relevance of mucin VNTR variation, we applied Locityper to genotype 4,637 cystic fibrosis patients from the Cystic Fibrosis Foundation where whole-genome sequencing (WGS) data has been generated^9^. These patients had been previously phenotyped using the Kulich normal residual mortality-adjusted (KNoRMA) score from the Cystic Fibrosis Gene Modifier Consortium^52^ (Supplementary Figure 7), which integrates multiple FEV1 (forced expiratory volume) measurements over time with adjustments for disease progression, mortality, age, sex, and *CFTR* mutations to create a standardized measure of lung disease severity. Higher KNoRMA scores indicate better lung function and less severe disease progression, while lower scores are associated with poor outcomes like increased mortality.

We tested four complementary association models incorporating sex and genetic ancestry as covariates (Supplementary Figures 8-9): continuous VNTR length, haplogroup dosage, binned length categories, and extreme allele comparisons (Methods). Given the variable genotyping accuracy observed across loci, we restricted analysis to mucins with ≥95% concordance for models requiring precise length estimates (*MUC5AC*, *MUC5B*, *MUC7*, *MUC17*, and *MUC21*; Supplementary Tables 6-9), while the haplogroup dosage model included all mucins except those with poor haplogroup-level accuracy (*MUC2*, *MUC4*, *MUC12*, and *MUC20*). After Bonferroni correction for multiple hypothesis testing, we identified a significant association between *MUC1* and cystic fibrosis severity (Figure 6c). Specifically, longer *MUC1* VNTR alleles in haplogroup H2 were associated with higher KNoRMA scores, indicating milder disease progression. This finding suggests that *MUC1* VNTR length variation is a likely modifier of cystic fibrosis lung disease severity, with longer alleles conferring a protective effect against disease progression.

## DISCUSSION

Mucins rank among the most structurally variable loci in the human genome, a feature shared with immune-related gene families subject to ongoing adaptive pressure^53^. The mucin gene family is well established as having substantial biological and clinical importance, yet its sequence architecture has been challenging to resolve with standard sequencing methods over the last 20 years^14,16,19^. We applied long-read sequencing and haplotype-resolved pangenome assemblies to create a high-resolution resource for the mucin and human genetics communities. We used the T2T-CHM13 reference^22^ to correct liftover errors in mucin gene models, producing a consensus set of full-length transcripts that faithfully represent the coding VNTRs, and then assembled mucin loci completely and accurately across diverse haplotypes^15,18^ (Figure 1). This shifts mucin genetic analysis from single-reference characterization to population-level inference. The resource we developed has additional utility beyond population genetics. Our curated protein annotations enable more sensitive peptide identification in mucin glycoproteomics and the complete protein sequences provide a foundation for systematic assessment of mucinase specificity across diverse alleles^54,55^ (Figure 3, Extended Data Figure 3, Supplementary Table 3). More broadly, this pangenome approach should accelerate analysis of other gene families where repetitive architectures encoding proteins have historically evaded standard tools^18^.

Despite their shared sequence features of repeat-rich, heavily glycosylated domains, the canonical mucins we characterized are strikingly heterogeneous in length, copy number, and protein structure. Each gene depicts a distinct pattern of polymorphism, repeat composition, and population genetic history (Figure 4), and these differences hold even between mucins mapping in close proximity (Figure 1). In general, secreted/gel-forming and tethered mucins possess distinct properties diverging not only in subcellular localization and assembly^5,56^ but also in genetic architecture, polymorphism profiles, and evolutionary trajectory. The tethered mucins span both extremes of size, from the comparatively compact *MUC21* to the massive *MUC12*, and both extremes of polymorphism, from the hypervariable *MUC4* to the nearly invariant *MUC16*. The four gel-forming mucins we annotated (*MUC2*, *MUC5AC*, *MUC5B*, and *MUC6*) cluster in the middle of both axes, occupying a comparatively narrow band of size and heterozygosity likely bounded by the structural constraints of their shared D-domain architecture^57^. These features suggest the mucins are better regarded as two distinct gene families in evolutionary, functional, and disease-association studies.

Within each class, several loci stand out. *MUC16* encodes one of the largest proteins in the human proteome yet resolves into only three distinct alleles differing only by 3 bp each, indicating selective constraint against length variation at this locus. Compared to other NHPs, this modal length appears to be specific to the human lineage. Conversely, its VNTR is also the most degenerate, indicating that sequence variation is less critical. Prior mammalian analyses have not identified *MUC16* as broadly constrained^58^, raising the possibility that the length constraint we observe is lineage specific. The functional basis for this is unknown, with potential ties to *MUC16*’s roles in epithelial barrier function and fertility, or its pathogenic role in ovarian cancer^59^.

Conversely, *MUC12*, *MUC1*, and *MUC4* sit at the opposite end of the diversity spectrum. The central VNTR-encoding exon of *MUC12* is, to our knowledge, the largest single protein-coding exon in the human genome in some humans (Figure 1). The combination of extreme size and limited prior characterization makes *MUC12* a strong candidate for future functional work. *MUC1* and *MUC4* are the most heterozygous loci we characterized and among the most variable genome wide. The chr3q29 region harboring *MUC4* and *MUC20* also warrants particular care for future studies due to higher-order structural variation in the region extending well beyond VNTR length^60^ (Figure 5). We identified 20 distinct long-range structural configurations that mediate *MUC20* copy number variability in ∼30% of haplotypes. This region is among the most variable in the human genome, making it perhaps unsurprising that *MUC4*, the most variable canonical mucin, sits within this larger landscape of structural variation.

Translating this haplotype catalog into disease association requires bridging long-read assemblies with short-read WGS cohorts that dominate existing resources, given that long-read assembly in large cohorts remains prohibitively expensive^61^. We pursued two complementary strategies (Figure 6), with the first mapping GWAS risk alleles onto our haplotype catalog to identify VNTR backgrounds that co-segregate with disease-associated variants. This approach generates candidate causal variants at otherwise inaccessible loci. Notable signals included short *MUC4* alleles linked to severe cystic fibrosis^9^ and long *MUC5AC* alleles linked to gastric cancer risk^62^. The second strategy directly genotypes mucin VNTRs in short-read data using Locityper^50^ with our expanded haplotype panel as reference, improving accuracy across most mucins. *MUC4* and *MUC12*, however, remain too complex for current short-read methods and will likely require new computational approaches designed for hypervariable multiallelic loci. SNP mapping is broad but indirect, whereas Locityper directly genotypes VNTRs but is limited to length variation captured by the reference panel. Used together, these methods offer a tractable path for testing mucin variation against disease in existing cohorts.

Applied to a cystic fibrosis (CF) cohort from the Cystic Fibrosis Whole Genome Sequencing Project, the Locityper framework identified long *MUC1* alleles as associated with milder lung disease, a signal that survived multiple testing correction (Figure 6c) and is supported by earlier orthogonal Southern blot validation from Guo et al. for *MUC1* haplotype length^63^. A parallel pattern emerged from SNP mapping at *MUC4*, where long alleles also appeared protective, though *MUC4*’s structural complexity prevented direct genotyping. The convergence of these signals across two tethered mucins suggests that longer tethered mucin VNTRs may yield a denser glycocalyx through additional glycosylation sites^64,65^ and a greater capacity for hydration, pathogen entrapment^66^, and inflammatory modulation^67,68^. Each of these features could be compensatory in a chronically inflamed CF airway prone to infection and dehydration^69^, a setting in which the burdens of *Pseudomonas aeruginosa* colonization and accelerated lung function decline are drivers of reduced life expectancy^70^. These clinical implications also extend to common CF therapeutics, where CFTR protein modulators like elexacaftor/texacaftor/ivacaftor (ETI) act in part by altering sputum mucin properties^71^ and where mucinases modulate MUC5AC and MUC5B protein levels^72^. Functional follow-up will be required to distinguish among these mechanisms, but the associations we describe here indicate that mucin genotyping could be a critical addition to the CF diagnostic odyssey.

More broadly, our results now establish the mucins and their hypervariable VNTRs as tractable targets for disease genetics. We provide a methodological pipeline for studying other repeat-rich, multiallelic loci that have similarly resisted standard approaches^13,16^. For each locus, we reconstructed haplotype phylogenies and defined tree-informed haplogroups that simplify VNTR diversity to increase power for disease association testing. The genetic architecture of complex epithelial disease is unlikely to be fully explained without considering the structural variation in mucins and the missing heritability therein^73^. By resolving variation that has remained invisible to GWAS for two decades, long-read pangenomes set the stage for the next phase of human disease genetics—one in which repeat-rich loci are no longer excluded but embraced.

## METHODS

### Data sources and QC

Human WGS assemblies constructed with long-read sequencing were used for analysis of canonical mucin loci (n = 296 genomes), all of which are consented for open access with no data use restrictions. The majority of assemblies were obtained from the HPRC release 2 (n = 231) ^74^ and the HGSVC phase 3 (n = 60)^17^. An additional 5 genomes were obtained from HPRC release 1^75^ (n = 1 genome) and HGSVC phase 2 (n = 4, assembled by Plender et al.^19^). Sequencing statistics for the samples are available in the methods of the associated publications and assembly algorithm information is available in Supplementary Table 2. Additionally, NHP genomes from eight species were used for comparative analyses. Those assembled by Yoo et al.^41^ include chimpanzee, bonobo, gorilla, Sumatran orangutan, Bornean orangutan, and Siamang gibbon (n = 5 genomes, one per species). Regional assembly for all ape haplotypes (human and NHP) was assessed using Flagger v0.3.3. The macaque was assembled by Zhang et al.^42^ and the marmoset was assembled by Hebbar et al.^43^ We performed manual curation with read alignment for these additional genomes.

The HPRC and HGSVC assemblies represent the same individuals as 1KGP samples and are classified into population and superpopulation labels based on recruitment geography or having at least three grandparents from the same continental region^34^. In total, these include 100 African (AFR), 52 Admixed American (AMR), 61 East Asian (EAS), 39 European (EUR), and 44 South Asian (SAS) genomes. We assessed the correlation of these annotations for HPRC release 2 haplotypes (n = 231) with local ancestry predictions per locus using Point Cloud Local Ancestry Inference (PCLAI), a method that groups alleles into four clusters that correspond to AFR ancestry, EUR ancestry, SAS ancestry, and EAS/AMR ancestry^35^. To more accurately reflect local ancestry at each mucin, we performed most downstream population comparisons using these ancestry clusters in HPRC haplotypes only (excluding recombination rate comparisons, see below).

### Gene model reconstruction and haplotype annotation

Loci chosen for inclusion in this study were determined based on mucin protein domain information^76^ and sufficient RefSeq/VNTR annotations in CHM13^22^ (Supplementary Table 1). Loci with gene models that did not span the complete VNTR sequence in CHM13 were manually curated to maintain ORFs consistent with GRCh38 annotations^77^. Human and NHP haplotypes were aligned to CHM13 using minimap2 v2.24^78^ with allowance for up to 20% sequence divergence, a minimum chaining score of 25000, and suppression of secondary alignments. Coordinates for mucin loci within haplotypes were extracted using rustybam v0.1.29^79^ and haplotype sequences were mined using seqtk v1.3^80^. Mucins with significant ancestry cluster stratification for VNTR exon lengths were determined via a Kruskal-Wallis test of variance (FDR adjusted, df = 4, α = 0.05) and pairwise comparisons were performed using Wilcoxon rank-sum tests (FDR adjusted, α = 0.05). Exons were translated into predicted proteins using Expasy (EMBOSS v.6.6.0^81^). VNTR exon sizes in NHP assemblies were estimated using a custom pipeline that mapped 500 bp flanking anchor sequences from the T2T-CHM13v2.0 reference to each assembly using minimap2. Soft-masked repetitive bases were trimmed from anchor ends prior to alignment, and only anchors with at least 100 bp of unique sequence were retained. For each mucin locus, the best upstream/downstream anchor pair mapping to the same contig and strand was identified, with locus-specific maximum span thresholds used to prevent paralogous mispairing. VNTR exon size was then calculated from the distance between paired anchors, including manual curation for overlapping anchors at smaller loci.

### Protein domain and repeat motif annotation

Protein domain annotations for the secreted, gel-forming mucins (*MUC2*, *MUC5AC*, *MUC5B*, and *MUC6*) were manually curated in a style consistent with Guo et al.^82^ and Plender et al.^19^ to identify variation in cysteine-rich and tandem repeat domain copy number. Tandem repeat protein motif variation was assessed across haplotypes for each mucin to reveal the most frequent motif, which was then compared to the canonical sequences determined by Chaturvedi et al.^23^ (excluding MUC21 and MUC22). Percent serine and threonine estimates were calculated based on total protein length in haplotypes that maintained complete ORFs. Mucin expression across epithelial tissues was assessed using data from the Genotype-Tissue Expression (GTEx) Consortium (v10)^83^, where median gene-level expression values (TPM) for epithelial tissue types were mapped for each mucin. Repeat motif degradation was quantified by aligning each repeat to the most frequent motif for each mucin (see above) and unit positional degeneration within the VNTR arrays was determined via an edge degeneration score. Repeat units were classified as “edge” (first and last N repeats, where N was adaptively determined as a fraction of the median repeat count) or “middle” and the score was determined as the middle mean identity minus edge mean identity (positive values indicate greater degeneration at array boundaries).

### Recombination rates, phylogenetic mapping, and haplogroup identification

Recombination rates (in cM/Mbp) in mucin regions were extracted from population-specific 1KGP recombination maps aligned to CHM13^40^. For each mucin locus, the mean recombination rate was calculated for five representative populations and compared to the genome-wide rates (computed via non-overlapping windows across the genome per population, then log10-transformed with a pseudocount (log10(rate + 1×10⁻⁴)) to normalize for the right-skewed distributions. A log-space z-score was computed for each mucin locus as (log10(query rate + 1×10⁻⁴) − log mean) / log standard deviation.

Phylogenetic trees per mucin locus were constructed using likely neutrally evolving sequences corresponding to non-repetitive intronic and intergenic sequences from human haplotypes that passed Flagger QC and two chimpanzee haplotypes for outgrouping. Global pairwise alignment was conducted using MAFFT v7.487^84^ (100 iterations). Alignments were inspected using Jalview v.9.0.5^85^ and erroneous regions were manually removed. Iqtree v.1.6.12^86^ and ggtree v.3.2.1^87^ were used to construct and visualize maximum likelihood trees based on 1,000 bootstraps.

Haplogroups were inferred independently per mucin based on a two-stage pipeline. First, patristic distances for all haplotype tips were computed from the rooted phylogenetic trees and subjected to hierarchical clustering (UPGMA). Then, a range of tree height thresholds were scanned, and at each threshold, candidate clusters were retained only if they contained more than three haplotypes and were supported by at least 70 bootstraps at their most recent common ancestor (MRCA) node. The optimal threshold was selected by maximizing the squared correlation between cluster assignment and VNTR length (one-way ANOVA), thereby yielding haplogroup definitions that best capture VNTR length variation in a phylogenetically supported manner. Orphan tips that failed initial clustering were reassigned to the nearest surviving cluster based on patristic distance. Full sequence trees were then pruned to the MRCA nodes of haplogroups and branch lengths were calibrated by scaling the root to 6.4 mya (estimated human–chimpanzee divergence^88^).

### Exploration of structural variation in chr3q29 and *MUC20* gene conversion

Structural configurations of chr3q29 across the *MUC4/MUC20* locus (chr3:198,100,000-198,800,000 in CHM13) were determined based on major inversion status across the region, *MUC4* partial gene duplication (number and length) with associated inversions, and *MUC20* partial or whole gene duplications (number and length) with associated inversions. Haplogroups were visualized using SVbyEye v0.99.0^89^ and annotated with DupMasker^90^ segmental duplication tracks (v4.2.1-p1). For haplotypes with full duplications of *MUC20*, the primary copy of the gene was determined via proximity to *MUC4* (closest = main designation, any additional = extra). The *MUC20* gene copy tree was constructed with all non-repetitive sequences (coding and noncoding, excluding the VNTR exon, total ∼19 kbp) in the same manner as the other phylogenetic trees (see above). Gene conversion between major *MUC20* copies and additional full copies was assessed using three tools in openRDP^91^ v0.1.0 in python v3.10.6^92^. The *MUC20* alignment was subsampled to 30 representative sequences along the tree due to the large number of haplotypes, including 2 chimpanzee *MUC20* haplotypes, 8 extra human copies of *MUC20* selected by greedy furthest-point sampling from all available extra copies (maximizing pairwise nucleotide distance), and 20 main copies from the same individuals in the eight extras and additionally diverse main copies. The sequences were trimmed of leading gaps and analyzed for gene conversion signatures using GENECONV^46^, Bootscan^47^, and 3Seq^47^. GENECONV identifies tracts of shared polymorphic sites between pairs that exceed expectations due to chance, Bootscan uses a sliding-window phylogenetic approach with bootstraps to detect topology switching (i.e., conversion breakpoints), and 3Seq evaluates triplets for nonrandom clustering of parental sites using an exact nonparametric test (p<0.05).

### GWAS risk allele assignment to mucin haplotypes

GWAS catalogue SNP associations were retrieved for each mucin using the NHGRI-EBI GWAS Catalog REST API^93^ and were parsed to extract reference SNP cluster IDs (rsIDs), reported traits/phenotypes, EFO-mapped trait terms, risk alleles, p-values, and effect sizes. SNPs without all data were excluded from further analyses. rsIDs were queried in NCBI’s dbSNP API for genomic coordinates in GRCh38. Given that rsIDs are not fully mapped to CHM13, SNP positions on HPRC/HGSVC haplotypes were determined by mapping a 100 bp window centered on each variant (+/-50 bp) from GRCh38 to CHM13 and using subsequent liftover in the same manner as gene model mapping (see above). VNTR length associations were assessed per SNP allele (major vs. minor allele) using Wilcoxon rank-sum tests with FDR adjustment.

### Genotyping using an expanded reference set with Locityper

Locityper v1.4.5^50^ was used to perform mucin VNTR and chr3q29 genotyping (inversion status, *MUC20* copy number). Reference alleles from the HPRC/HGSVC haplotypes were chosen per locus based on Flagger validation and availability of paired high-coverage (∼30×) short-read data through the 1KGP. Genotype concordance was assessed at the haplotype and haplogroup levels through a leave-one-out approach, where haplotype concordance denoted consistency for VNTR exon length per haplotype and haplogroup denoted phylogenetic clade prediction. To assess genotyping accuracy and estimate the panel size required for reliable mucin genotyping, we performed a subsampling analysis. In increments of 25 up to the full panel of 276 genomes, a random subset of genomes was drawn, and leave-one-out genotyping concordance was assessed per mucin, comparing predicted haplotype length and haplogroup against the known calls. A saturating curve was fitted to the resulting concordance-versus-panel-size data and a logistic model (concordance = A / (1 + exp(−k(n − n₀)))) was used to extrapolate the number of genomes required to reach 95% concordance for both VNTR length and haplogroup independently, with A as the asymptomatic maximum, k as the growth rate, and n₀ as the inflection point. This model was more biologically appropriate for the concordance metric than a log-linear or power law model because it saturates at a ceiling and fits the observed data.

We additionally evaluated the effect of region-specific sequence weighting, a feature introduced in Locityper v0.17 that allows differential weighting of genomic intervals genotyping. Five weighting schemes were tested: (1) no weights (standard Locityper); (2) down-weighted flanks, where intronic and exonic sequences were assigned intermediate weights (introns: 0.2, exons: 0.1) and the VNTR was set to 1, while flanking sequence upstream of the start codon and downstream of the stop codon was set to 0.01; (3) down-weighted VNTR, where all non-VNTR sequence was weighted at 1 and the VNTR was set to 0.5; (4) LD-informed weighting, where the VNTR was set to 1, sequences in the same superpopulation-specific LD block as the VNTR were set to 0.5, other coding and intronic sequence to 0.1, and flanks to 0.01; and (5) up-weighted flanks, where flanks and introns were set to 0.1, exons to 0.2, and the VNTR to 1. Genotyping concordance was compared across conditions to identify the weighting scheme that best recovered known VNTR length and haplogroup assignments from leave-one-out.

### Associations of mucin genotypes with severe cystic fibrosis

Locityper v1.4.5^50^ was used to genotype 4,637 short-read WGS from patients diagnosed with cystic fibrosis. These patient samples are part of the Cystic Fibrosis Foundation data repository and were previously phenotyped using the kulich normal residual mortality adjusted for CF-related mortality score (KNoRMA) from the Cystic Fibrosis Gene Modifier Consortium (GMC)^9,52^. Higher KNoRMA scores are indicative of better lung function and generally less severe disease. The *CFTR* genotype is included in the KNoRMA score, with most samples harboring the common F508del mutation (∼65%). Genetic ancestry principal component analysis (PCA) and sex were derived using Somalier^94^. Somalier was run once across all samples to ensure consistent PCA space and sites chosen for projection consist of high-frequency single-nucleotide polymorphisms (SNPs) from gnomAD exomes (n sites = 17,766). Ancestry inference was then projected against the 1KGP reference panel^95^ PCA space for the five superpopulations (AFR, AMR, EAS, EUR, and SAS). Principal components (PCs) 1-3 were retained as covariates to control for population stratification in all subsequent association testing. Sex was inferred from Y chromosome dosage using Somalier and included as a covariate in all models. Age is included in the KNoRMA score and therefore was not included as a covariate.

Four complementary models were run for testing mucin genotype associations with KNoRMA continuous phenotype scores (Supplementary Tables 6-9): (1) a continuous VNTR length model, where total diploid VNTR length was z-score scaled and used as a continuous predictor; (2) a haplogroup dosage model to test haplogroup-specific lineages rather than raw length; (3) a binned length dosage model to reduce dimensionality (alleles binned into short, medium, and long categories based on per-mucin tertile distributions); and (4) an extreme allele comparison where only individuals homozygous for the shortest or longest alleles (defined by median split) were tested. Based on Locityper haplotype leave-one-out concordances below a 95% accuracy threshold, only *MUC5AC*, *MUC5B*, *MUC7*, *MUC17*, and *MUC21* were included in models 1, 3, and 4; however, all mucins except *MUC2*, *MUC4*, *MUC12*, and *MUC20* were included in model 2 (four excluded mucins less than 95% haplogroup concordance) and was therefore the primary model considered. Bonferroni and FDR correction (Benjamini-Hochberg) were applied across mucin loci within each model independently.

### Use of Claude for data visualization and copy editing

Claude (Anthropic, claude.ai, Sonnet 4.6/Opus 4.7) was used for coding assistance and general data visualization in R v4.3.3^96^. No AI tools were used to analyze any human genotype/phenotype data directly. All content was reviewed and verified by the authors, who take full responsibility for the accuracy of the final manuscript.

## DATA AVAILABILITY

Cystic fibrosis WGS data used in this study are available upon request through the Cystic Fibrosis Foundation Whole-Genome Sequencing project at. Restrictions on access to data are to ensure patient privacy for all persons in the WGS database. We thank the patients who participated in this cohort for their generosity in supporting this research.

## CODE AVAILABILITY

Custom scripts used for analysis in this study are publicly available at https://github.com/plendere/mucin_pangenome.

## AUTHOR CONTRIBUTIONS

E.G.P. and E.E.E. conceived and planned the experiments. W.K.O. and J.D.B. provided input for project design. General analysis and methods development was conceived and performed by E.G.P., J.L., and E.E.E. T.P. and T.M. provided support for E.G.P. with Locityper analyses. I.W. & J.W. assisted in Locityper implementation. W.W.G. and M.J.B. facilitated CF WGS data transfer to E.G.P. K.M.M. provided technical and scientific consultation. All data visualization was designed by E.G.P. and E.E.E. E.G.P. and E.E.E. wrote the manuscript, with edits and reviews from all authors.

## ACKNOWLEDGEMENTS

We thank Tonia Brown for assistance in editing this manuscript and Devin Schweppe, Brian Browning, and Nick Riley for their intellectual contributions to the analysis. The human cell lines used in the assemblies analyzed here were originally obtained from the NIGMS Human Genetic Cell Repository at the Coriell Institute for Medical Research (Supplementary Table 2) and were part of the 1000 Genomes project. These data are consented for open access with no use restrictions. Research reported in this publication was supported, in part, by the U.S. National Human Genome Research Institute of the National Institutes of Health (NIH) under grants HG002385, HG010169, and HG007497 to E.E.E and HG013748 to T.M. The content is solely the responsibility of the authors and does not necessarily represent the official views of the NIH. E.E.E and J.D.B. are investigators of the Howard Hughes Medical Institute.

This article is subject to HHMI’s Immediate Access to Research policy, which requires that this article be made publicly available as initial and revised preprints deposited on a designated preprint server under a CC BY 4.0 license.

## ETHICS DECLARATIONS

E.E.E. is a scientific advisory board (SAB) member of Variant Bio, Inc. J.D.B. is on the scientific advisory boards of Apriori Bio and the Vaccine Company. J.D.B. consults for GlaxoSmithKline and Pfizer. All other authors declare no competing interests.

## EXTENDED DATA

**Extended Data Figure 1.**
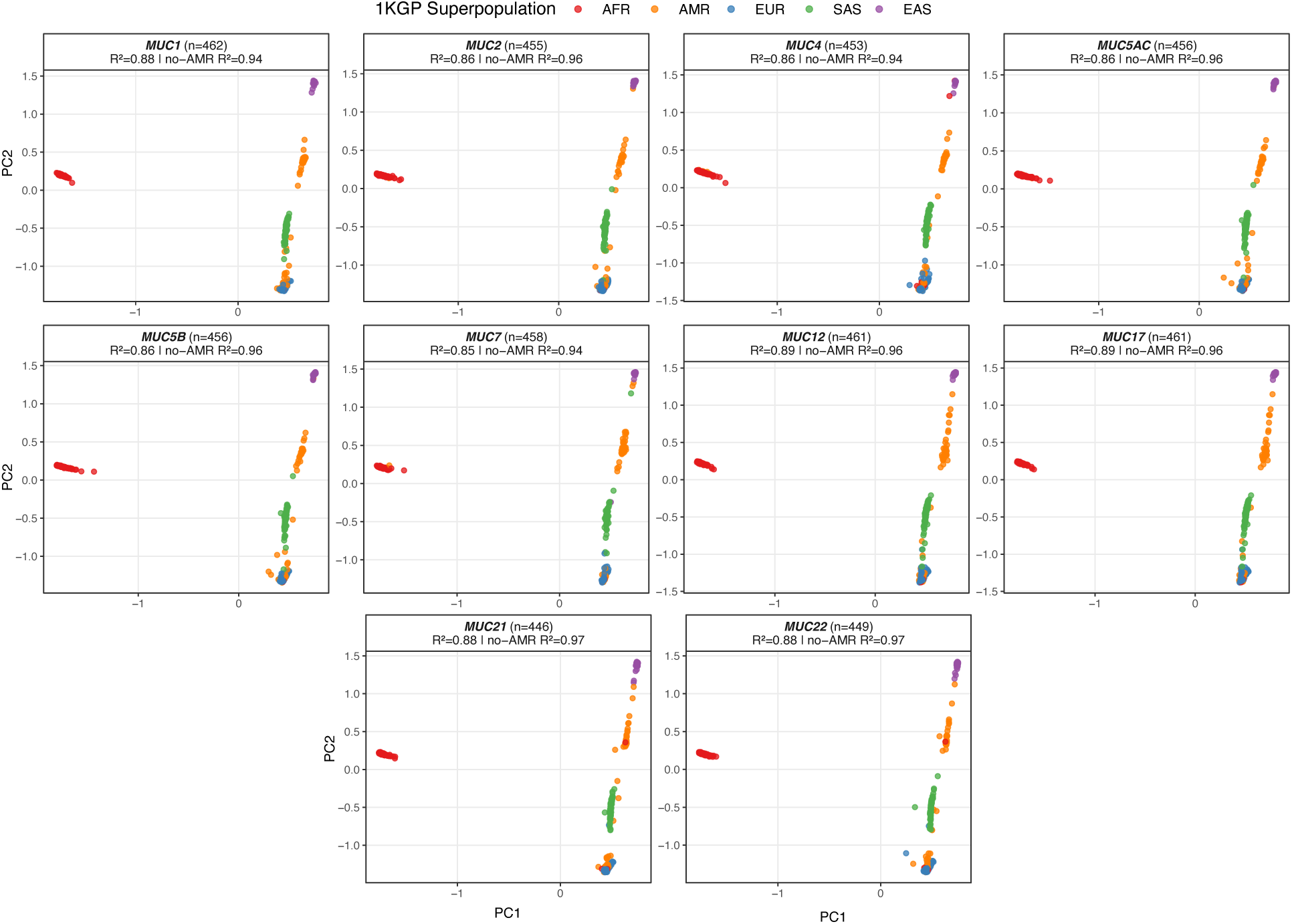
PCLAI clustering correlation with 1KGP superpopulation labels in HPRC haplotypes. Per-locus principal component (PC) coordinates (PC1, PC2) from PCLAI (Principal Cloud Local Ancestry Inference) for each HPRC haplotype, faceted by mucin gene. PCs were averaged across all PCLAI windows overlapping the locus and points are colored by 1KGP superpopulation assignment. Correlation performed using PERMANOVA R2 (1000 permutations, Euclidean distance) for all five superpopulations included and after excluding AMR.

**Extended Data Figure 2.**
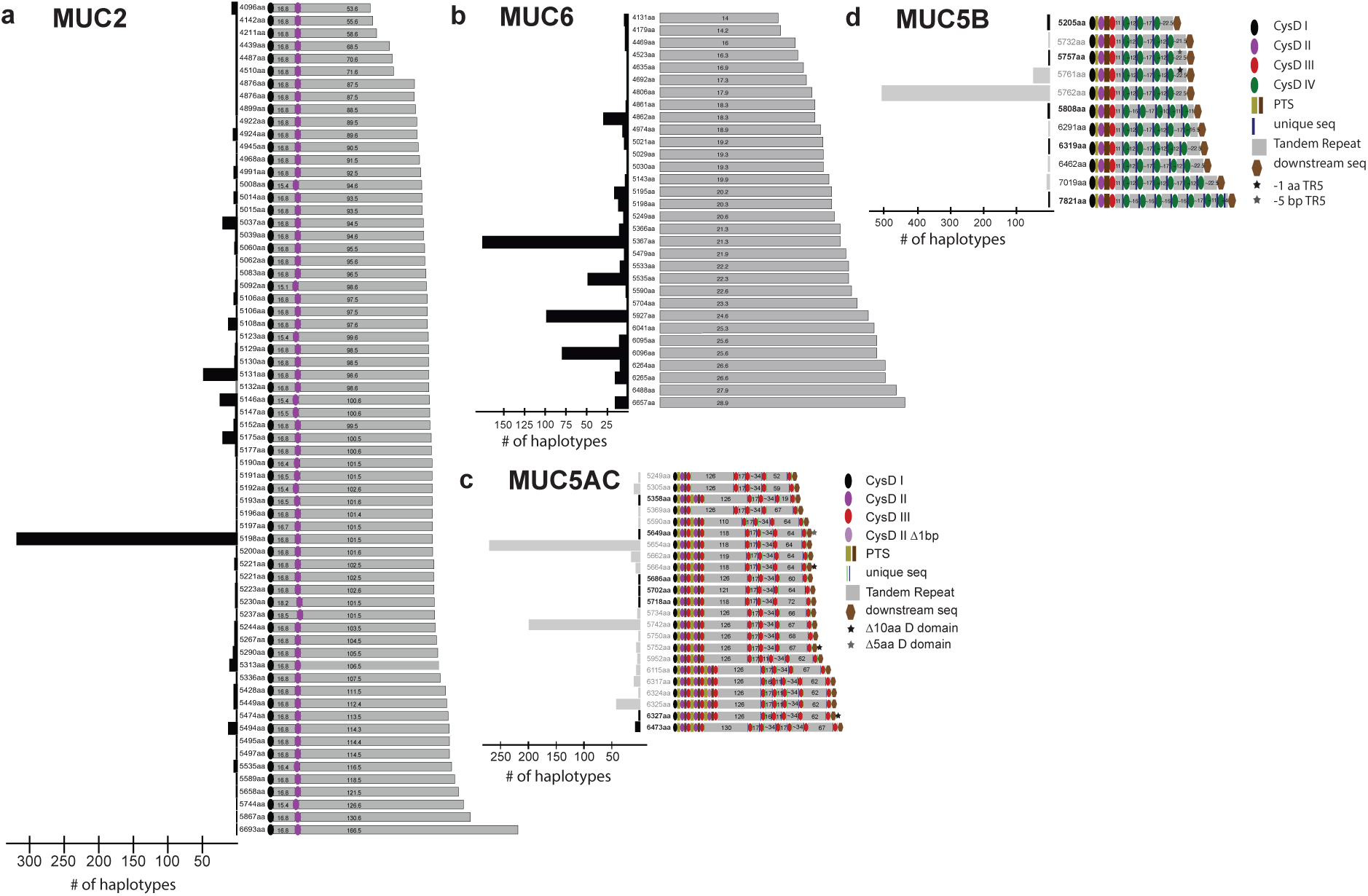
Protein domain diagrams for the secreted, gel-forming mucin loci. **a-d,** Diagrams represent domain annotations modeled from Guo et al.^82^ and Plender et al.^19^ CysD corresponds to cysteine-rich domains and PTS corresponds to proline-, serine-, and threonine-rich domains.

**Extended Data Figure 3.**
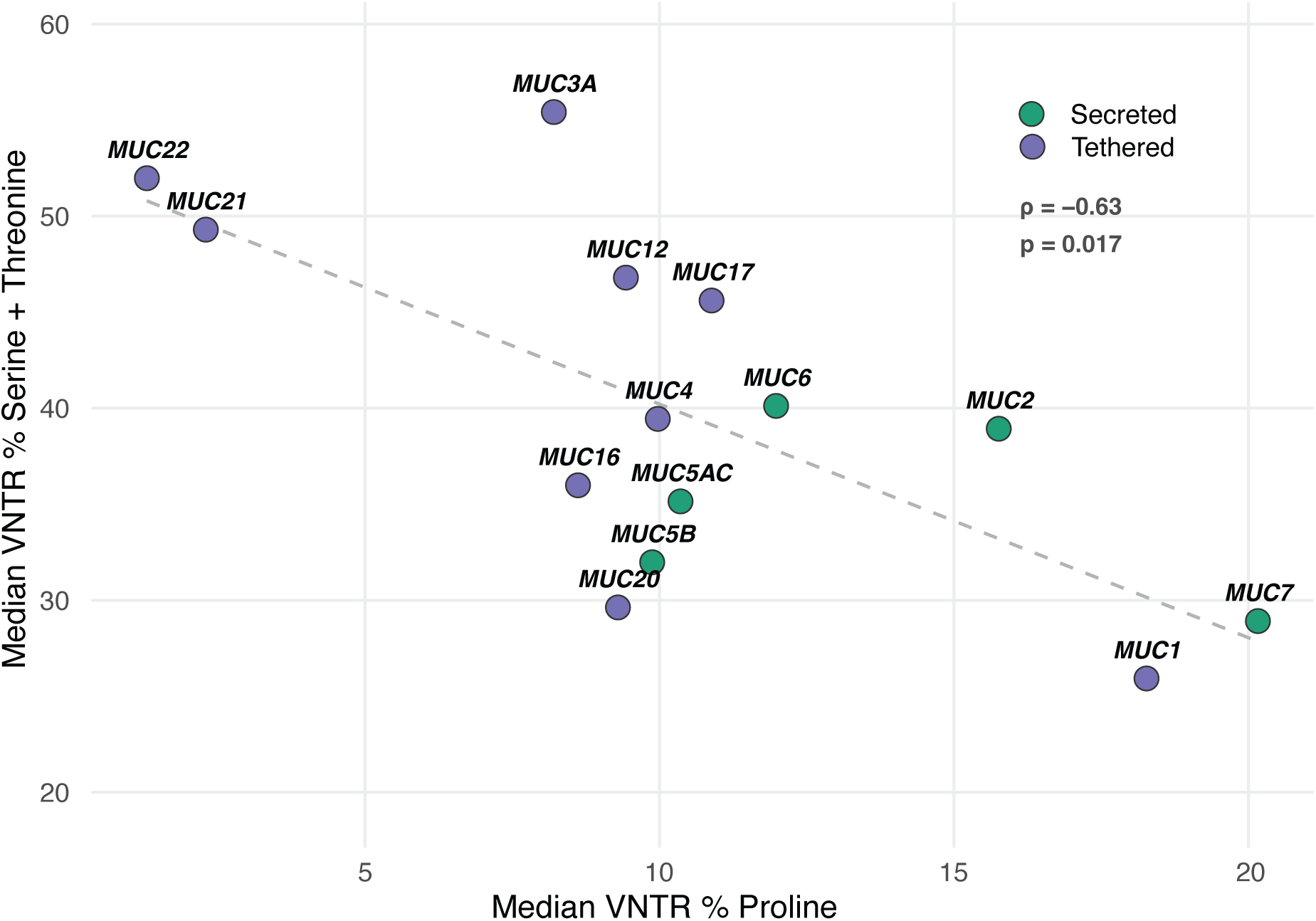
Correlation analysis of proline vs. serine/threonine abundance in mucin VNTRs. Correlation between median haplotype % S/T in the VNTR relative to median haplotype % proline in the VNTR was assessed using a Spearman rank correlation test.

**Extended Data Figure 4.**
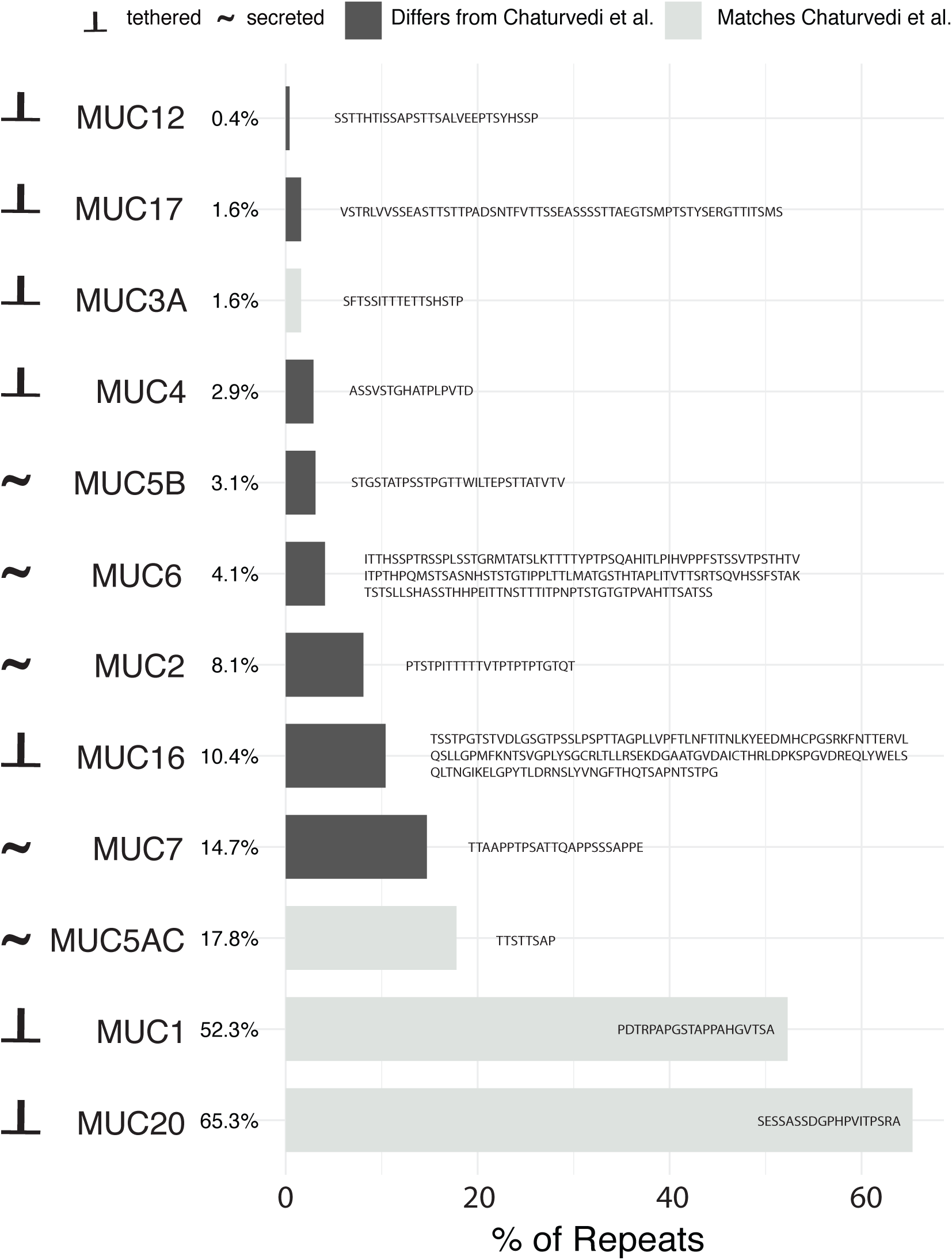
Comparison of reported canonical mucin amino acid motifs with Chaturvedi et al.^23^ and all HPRC/HGSVC haplotypes. Percentages represent the average frequency of the canonical repeat in the haplotypes. Updated canonical repeats reported to the left/overlayed against each bar per mucin protein.

**Extended Data Figure 5.**
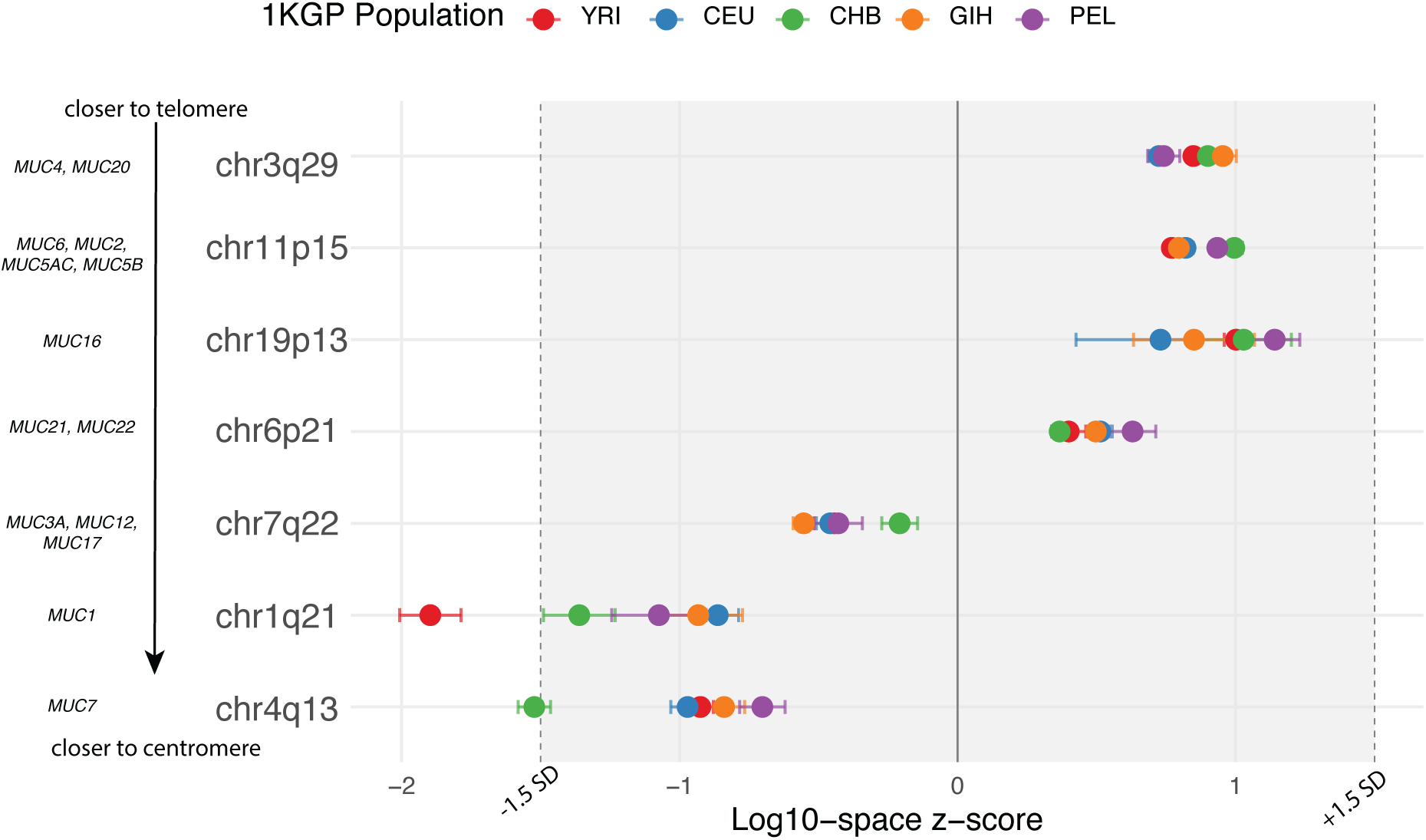
Recombination rates at mucin loci per continental representative populations. YRI = Yoruba in Nigeria, CEU = Utah residents from North and West Europe, CHB = Han Chinese in Beijing, GIH = Gujarati Indian in Houston, PEL = Peruvian in Lima.

**Extended Data Figure 6.**
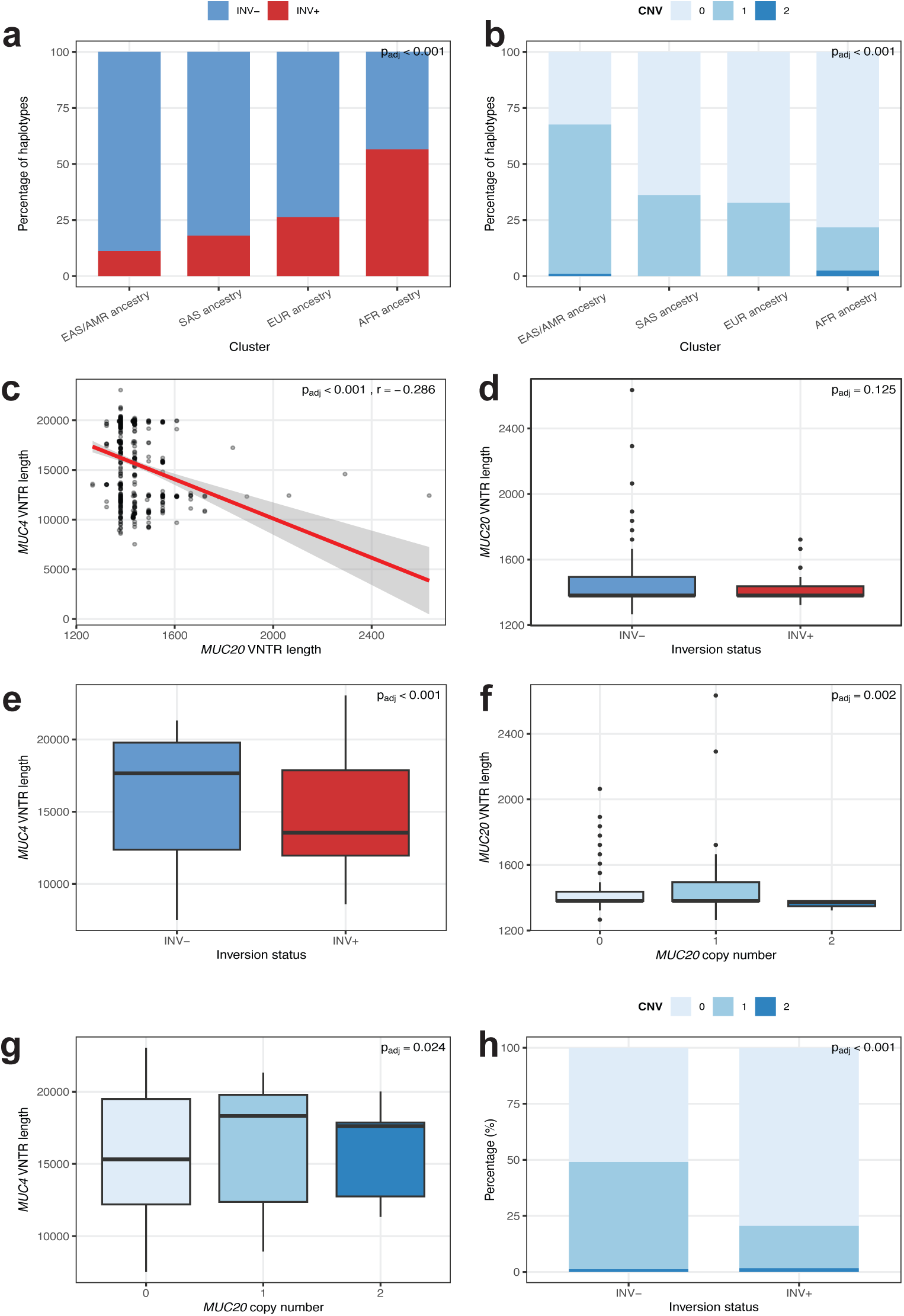
Correlation analyses for population genetic features in the chr3q29 region harboring *MUC4* and *MUC20*. **a,** Correlation of the inversion over *MUC4* and *MUC20* with ancestry cluster. Significance assessed by chi-squared test and Bonferroni corrected. **b,** Correlation of *MUC20* haplotype copy number and ancestry cluster. Significance assessed by chi-squared test and Bonferroni corrected (padj = p adjusted). **c,** Pearson correlation of *MUC4* VNTR length and *MUC20* VNTR length. **d,** Correlation of *MUC20* VNTR length and inversion status. Significance assessed via Wilcoxon rank-sum test. **e,** Correlation of *MUC4* VNTR length and inversion status. Significance assessed via Wilcoxon rank-sum test. **f,** Correlation of *MUC20* VNTR length and *MUC20* haplotype copy number. Significance assessed via Kruskal-Wallis test. **g,** Correlation of *MUC4* VNTR length and *MUC20* haplotype copy number. Significance assessed via Kruskal-Wallis test. **h,** Correlation of *MUC20* haplotype copy number and inversion status. Correlation assessed via chi-squared test.

**Extended Data Figure 7.**
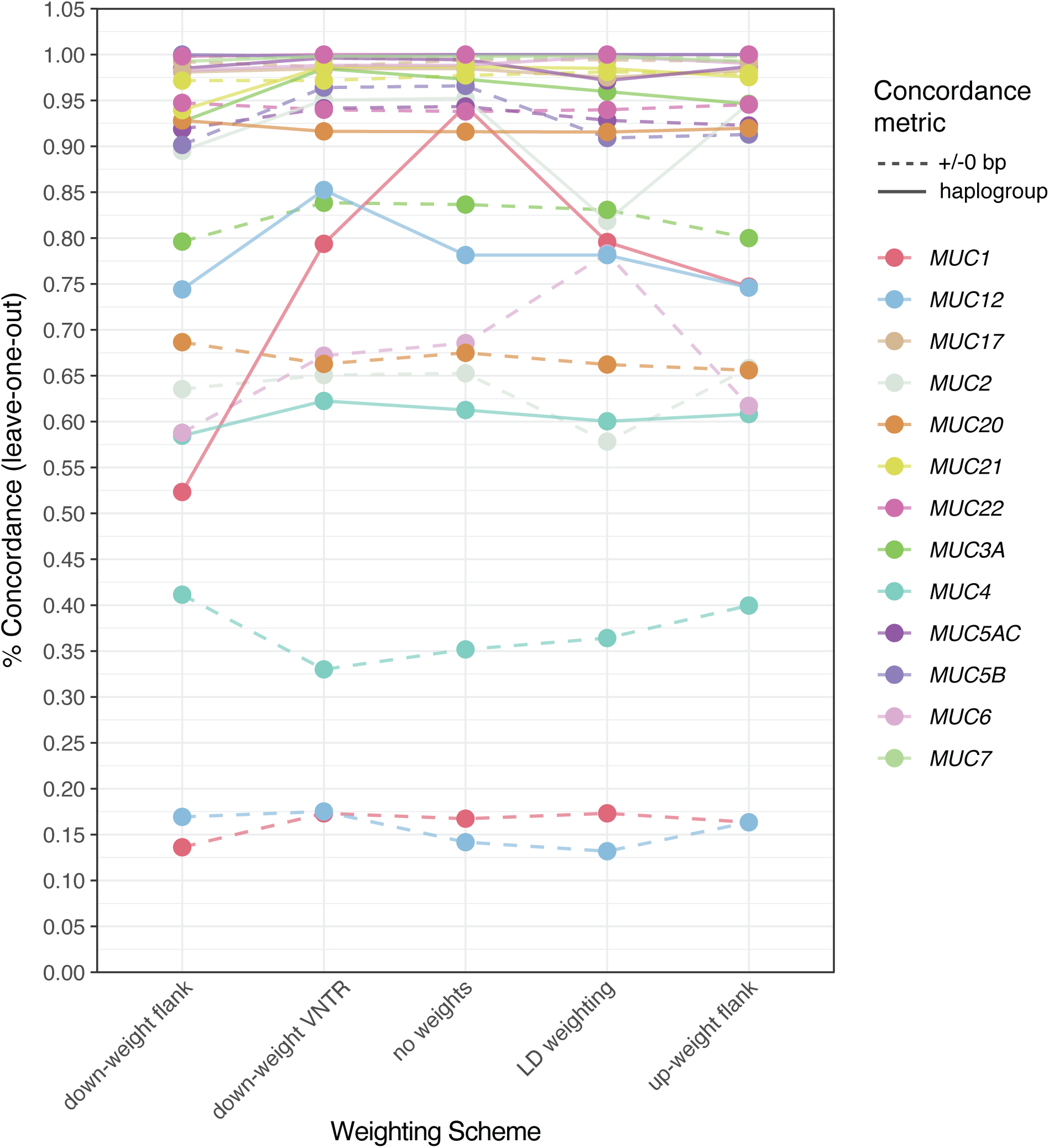
Testing of Locityper allele weighting schemes across mucin loci for haplogroup and haplotype level concordances in leave-one-out. Five weighting schemes tested were (1) no weights (standard Locityper); (2) down-weighted flanks, where intronic and exonic sequences were assigned intermediate weights (introns: 0.2, exons: 0.1) and the VNTR was set to 1, while flanking sequence upstream of the start codon and downstream of the stop codon was set to 0.01; (3) down-weighted VNTR, where all non-VNTR sequence was weighted at 1 and the VNTR was set to 0.5; (4) LD-informed weighting, where the VNTR was set to 1, sequences in the same superpopulation-specific LD block as the VNTR were set to 0.5, other coding and intronic sequence to 0.1, and flanks to 0.01; and (5) up-weighted flanks, where flanks and introns were set to 0.1, exons to 0.2, and the VNTR to 1.

**Extended Data Figure 8.**
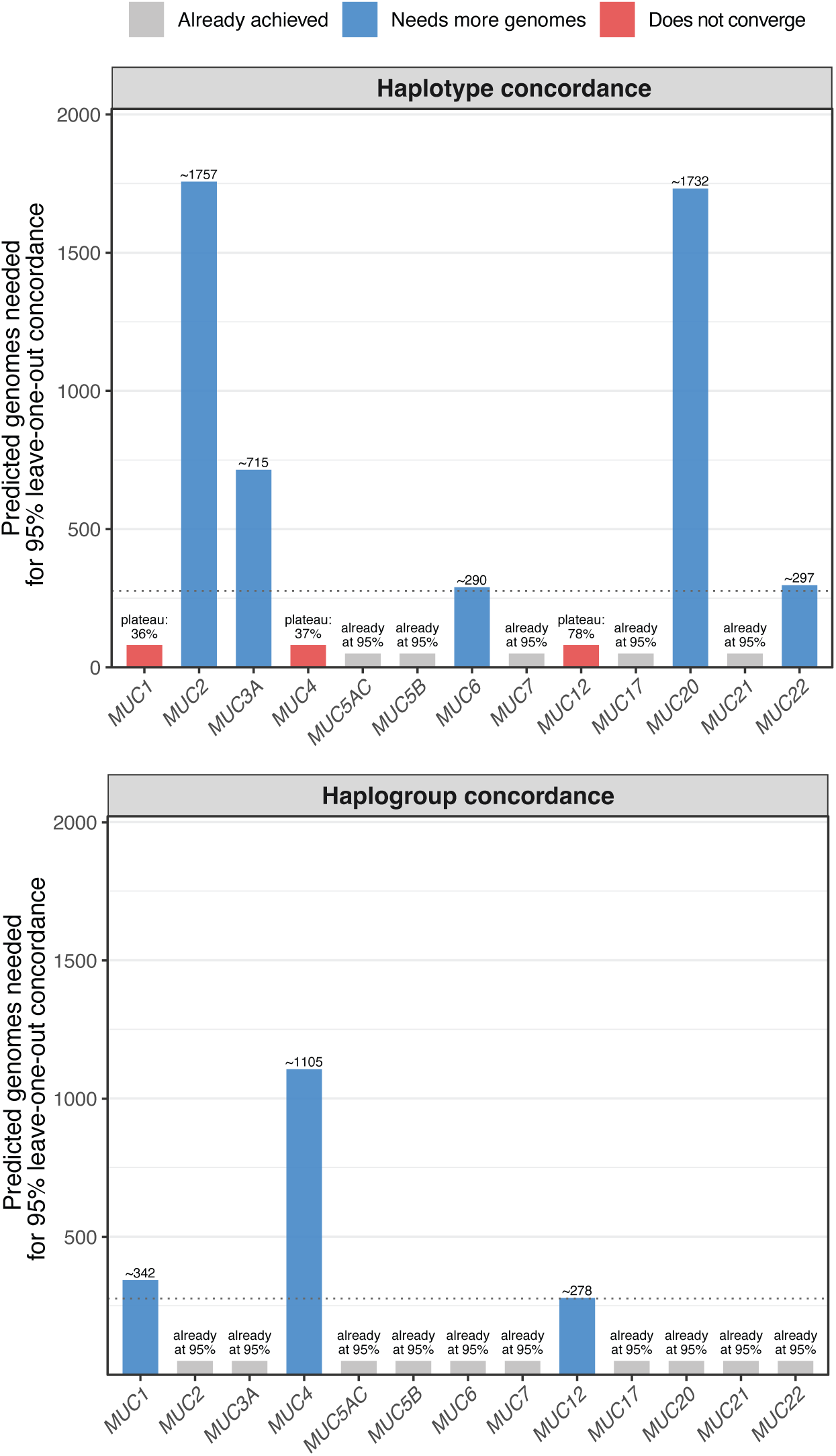
Predicted number of genomes needed to reach 95% genotyping concordance for Locityper per mucin. Estimation based on a logistic saturation model. Blue bars indicate mucins predicted to reach 95% concordance with additional genomes, with the estimated number shown. Red bars indicate mucins where the model asymptote falls below 95%, with the plateau percentage labeled. Gray bars indicate mucins that have already achieved 95% or higher concordance with the current panel (n = 276 genomes). The dashed line marks the current panel size.

## SUPPLEMENTARY INFORMATION

**Supplementary Figure 1.**
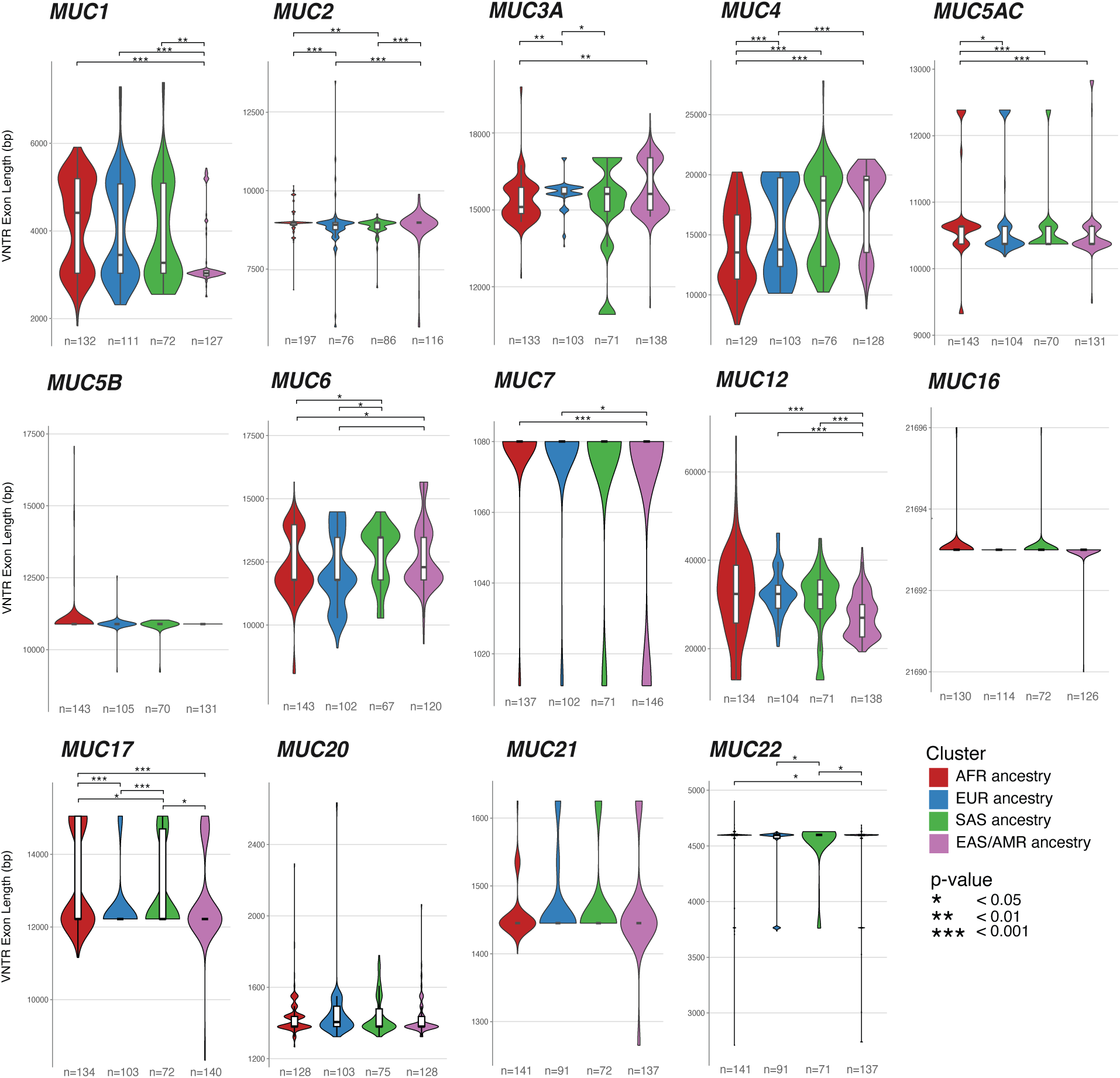
Mucin allele distributions with overlayed boxplots. P-values correspond to pairwise Wilcoxon rank-sum tests with multiple testing correction (FDR). n corresponds to number of haplotypes (HPRC).

**Supplementary Figure 2.**
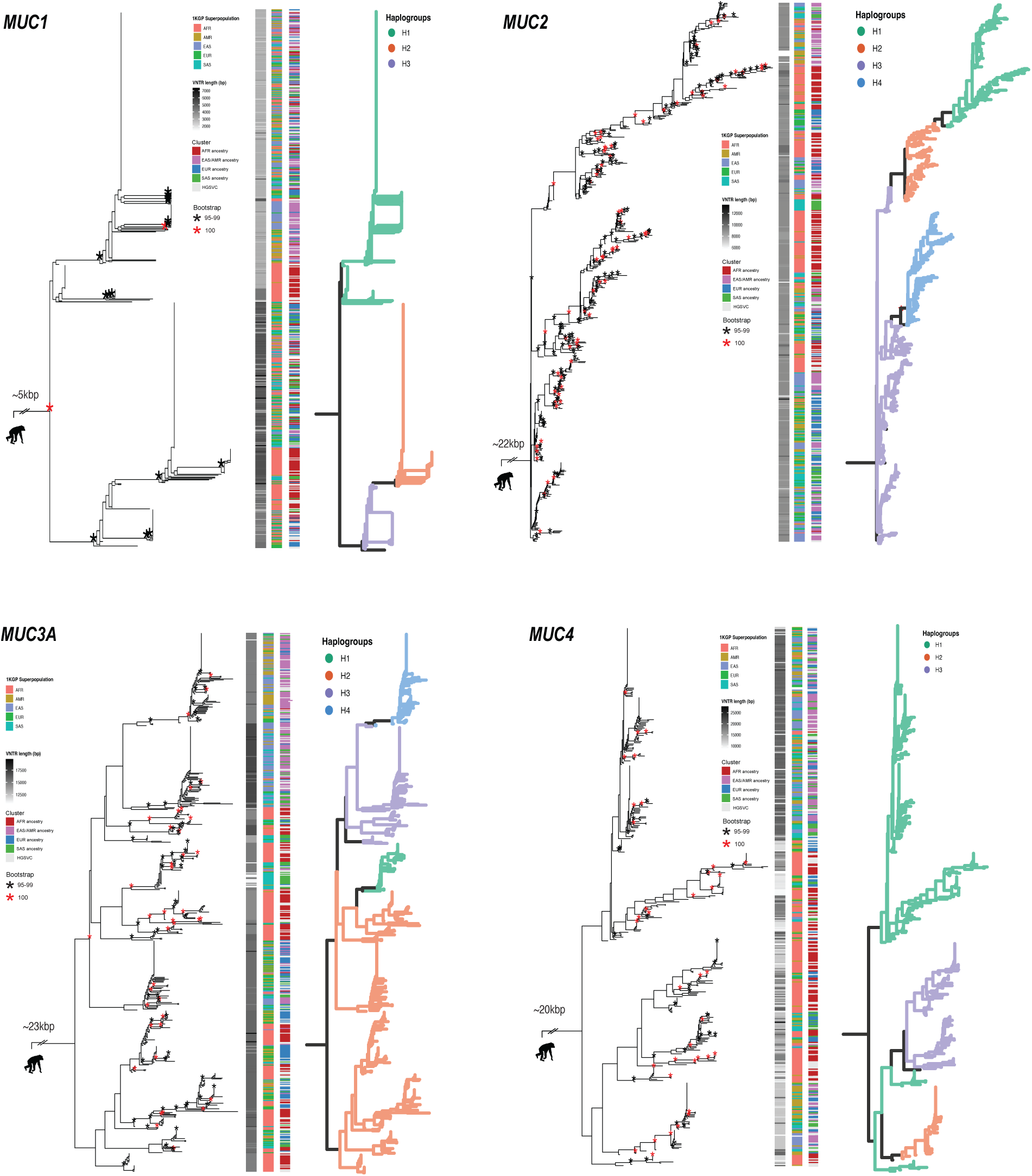
Complete phylogenetic trees and haplogroup overlay for *MUC1*, *MUC2*, *MUC3A*, and *MUC4*. Two chimpanzee haplotypes were used as outgroup.

**Supplementary Figure 3.**
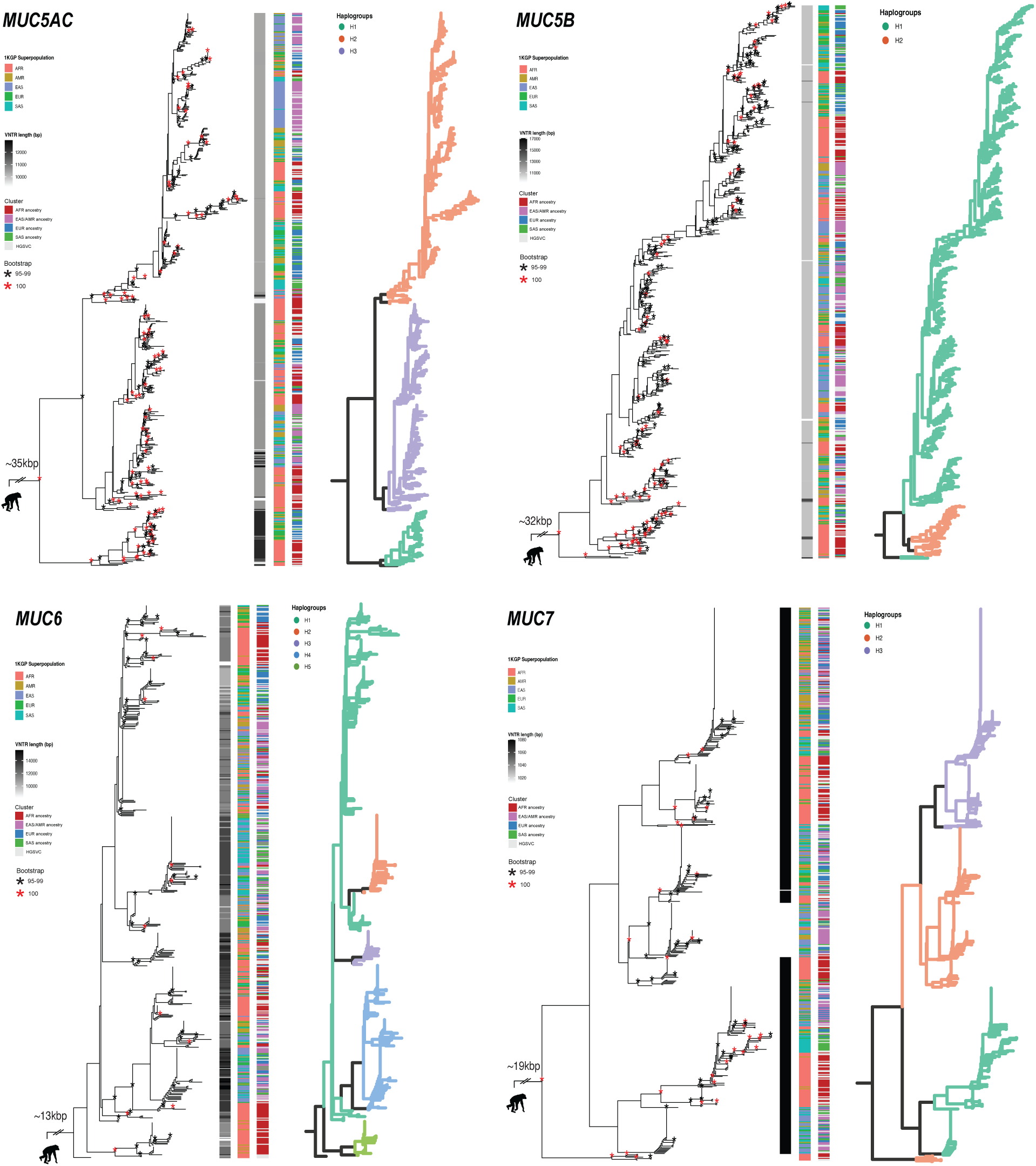
Complete phylogenetic trees and haplogroup overlay for *MUC5AC*, *MUC5B*, *MUC6*, and *MUC7*. Two chimpanzee haplotypes were used as outgroup.

**Supplementary Figure 4.**
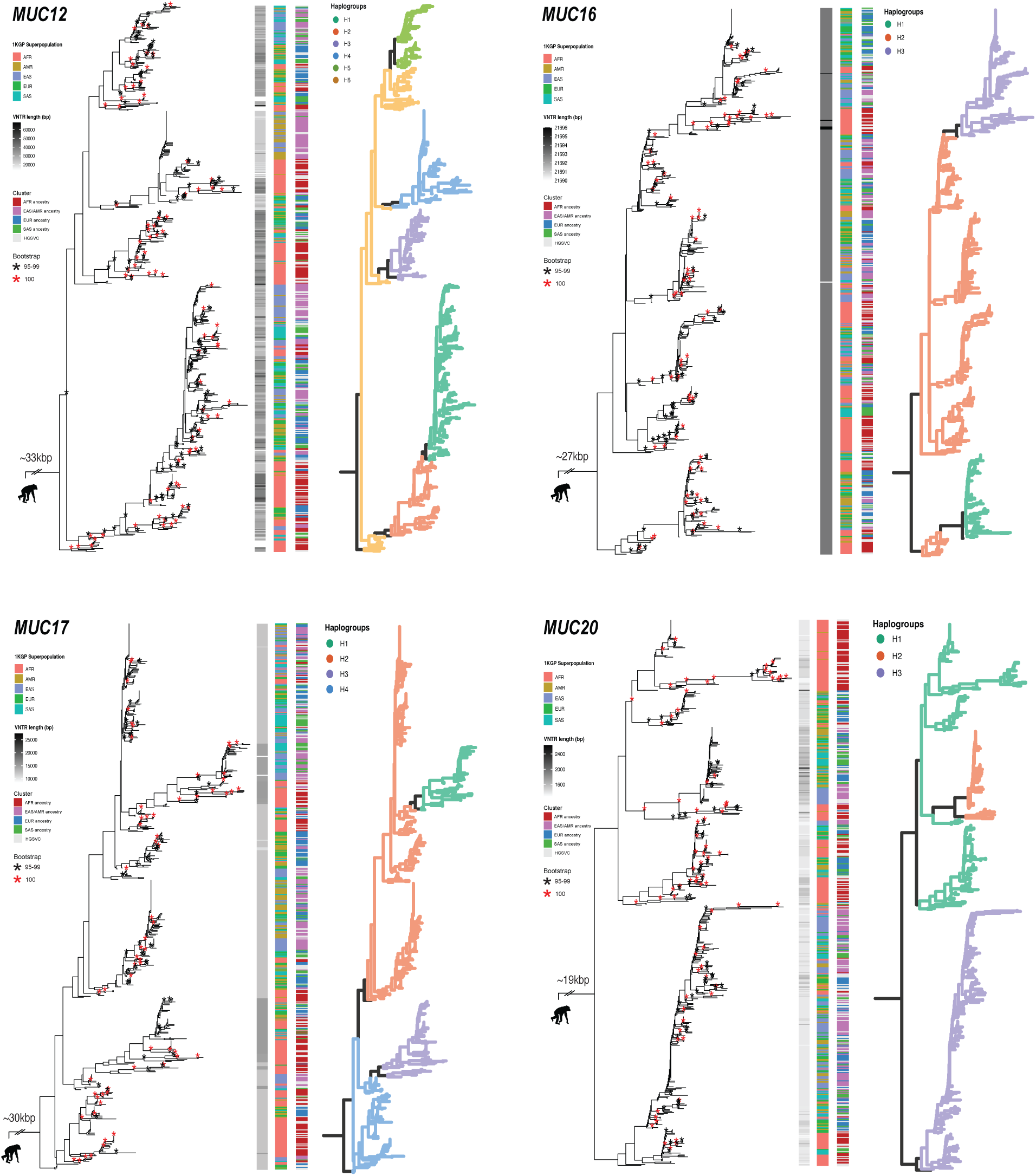
Complete phylogenetic trees and haplogroup overlay for *MUC12*, *MUC16*, *MUC17*, and *MUC20*. Two chimpanzee haplotypes were used as outgroup.

**Supplementary Figure 5.**
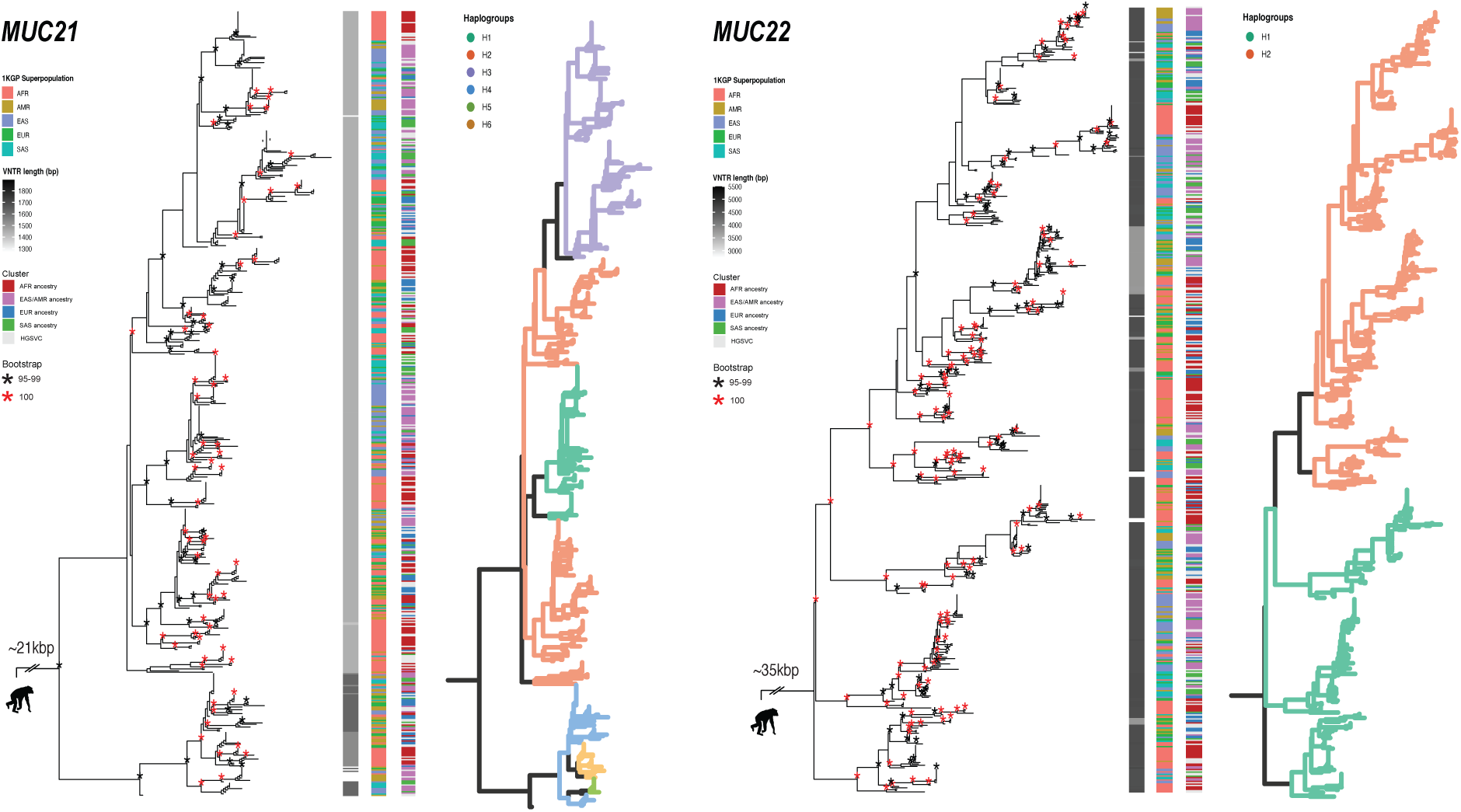
Complete phylogenetic trees and haplogroup overlay for *MUC21* and *MUC22*. Two chimpanzee haplotypes were used as outgroup.

**Supplementary Figure 6.**
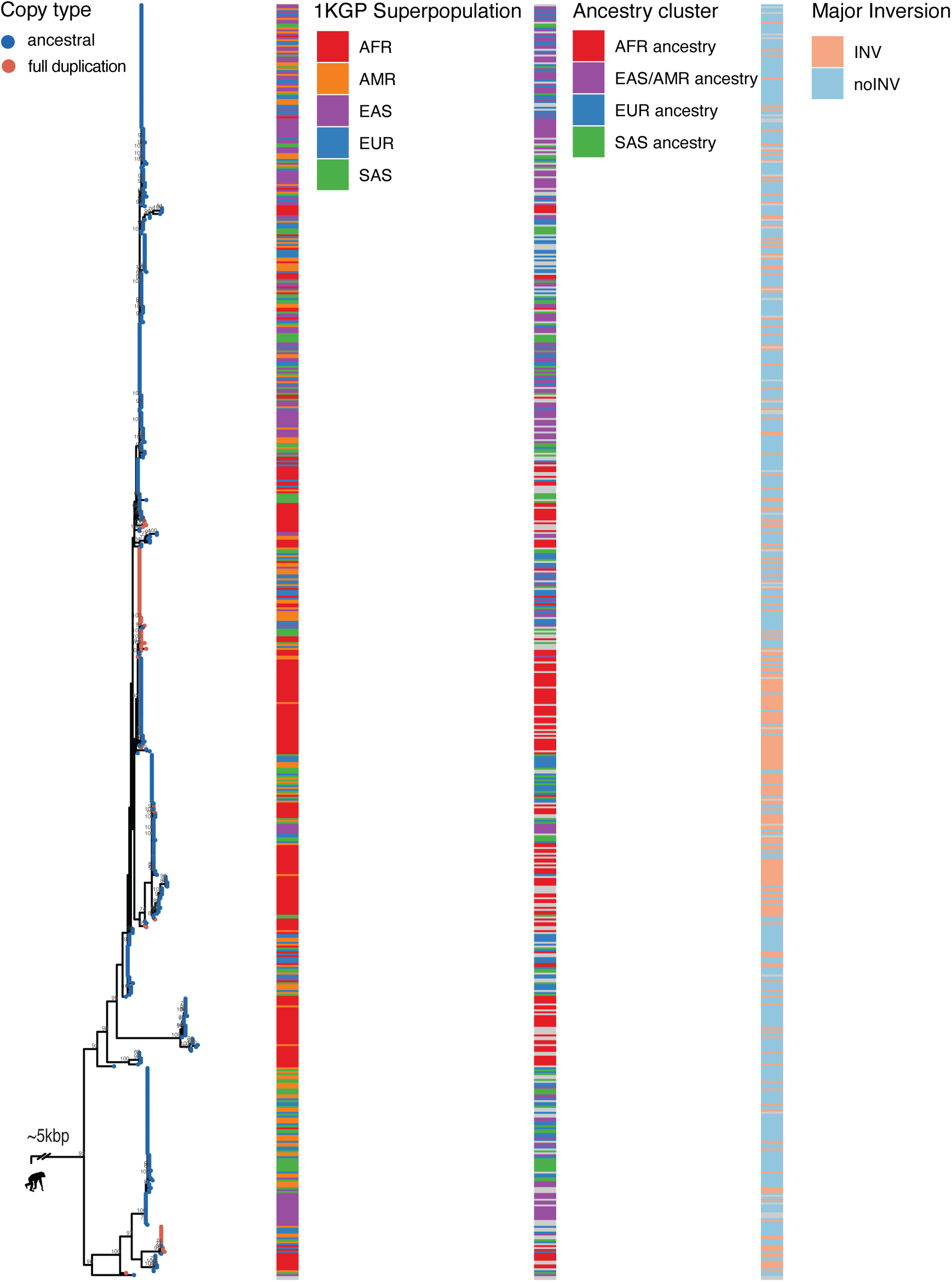
Phylogenetic tree of *MUC20* complete ancestral and full duplication copies in humans constructed without gene-converted sequence tracks. Two chimpanzee haplotypes used as outgroup. Numbers at nodes of tree correspond to bootstrap support. Gene conversion tracks determined via GENECONV results and removed from alignment before tree construction. Major inversion signifies presence or absence of inverted region overlapping with *MUC4* and *MUC20*.

**Supplementary Figure 7.**
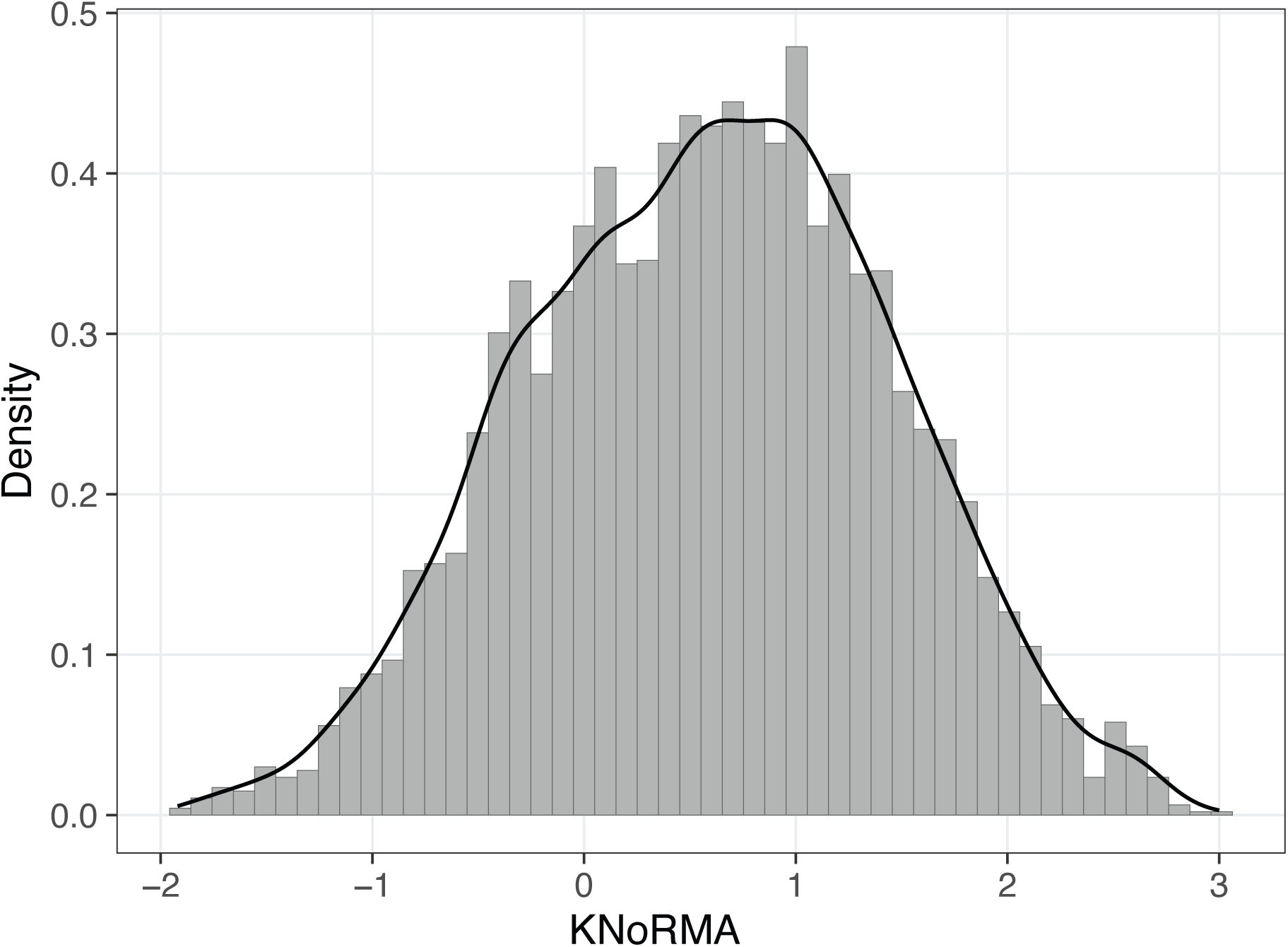
Distribution of KNoRMA scores across 4,637 paired short-read WGS data.

**Supplementary Figure 8.**
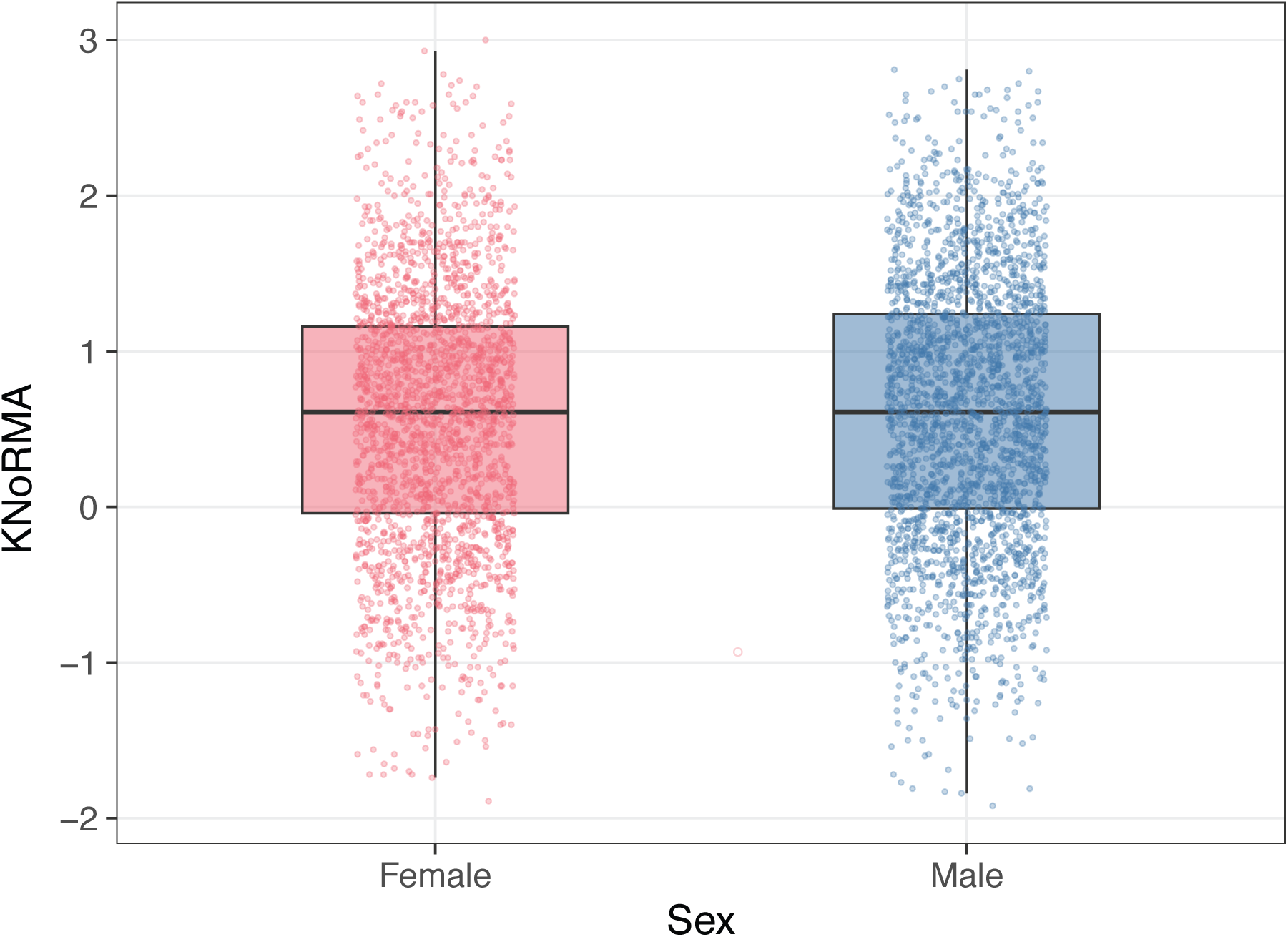
Sex distribution of KNoRMA scores across 4,637 paired short-read WGS data.

**Supplementary Figure 9.**
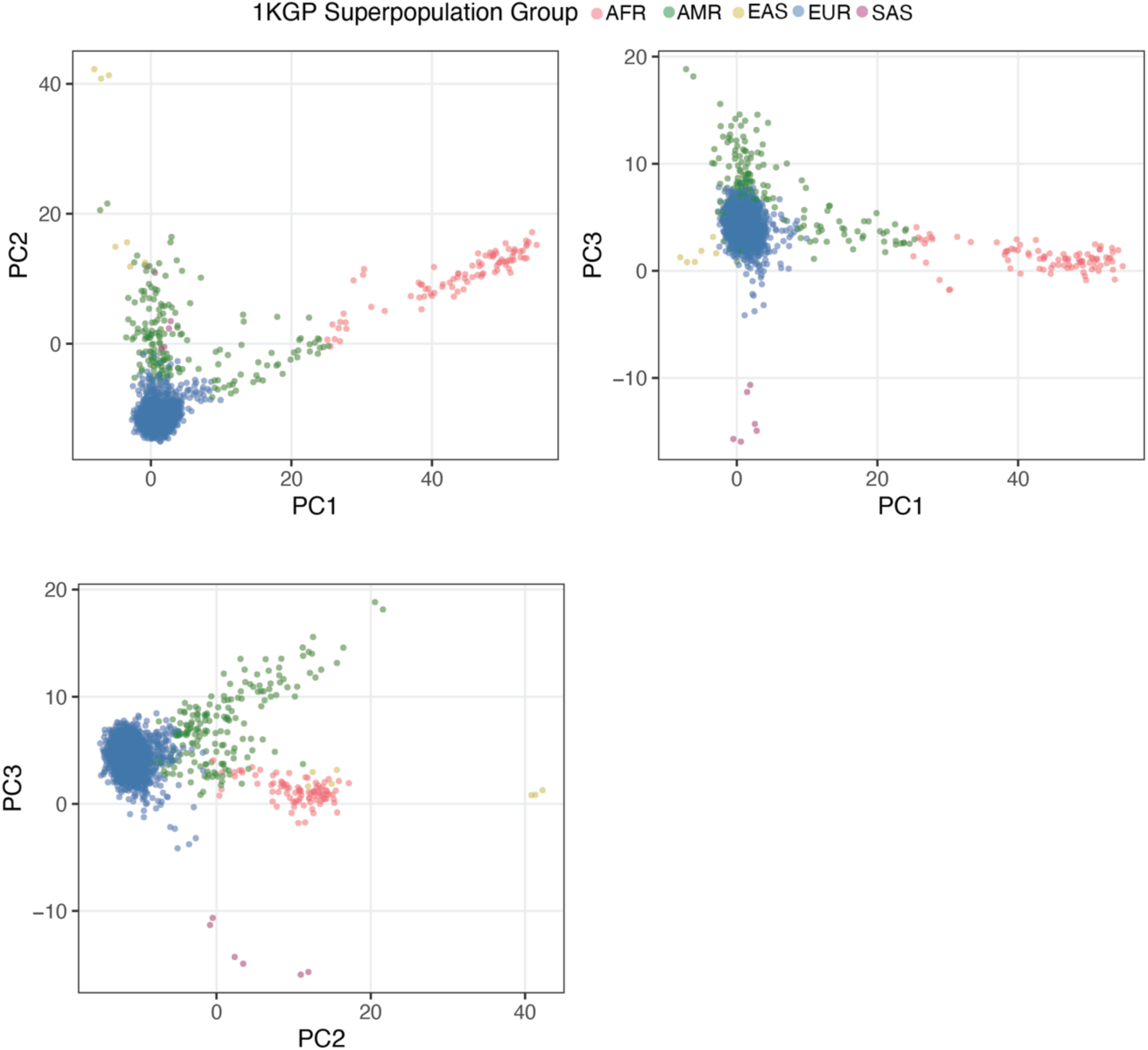
Principal components 1-3 from projection of 4,637 CF genomes onto 1KGP super-population labels.

